# Age and sex effects on blood retrotransposable element expression levels: Findings from the population-based Rhineland Study

**DOI:** 10.1101/2024.12.17.24319143

**Authors:** Valentina Talevi, Hang-Mao Lee, Dan Liu, Marc D. Beyer, Paolo Salomoni, Monique M.B. Breteler, N. Ahmad Aziz

## Abstract

Retrotransposable elements (RTEs) have been implicated in the pathogenesis of several age-associated diseases. Although model systems indicate that age- and sex-dependent loss of heterochromatin increases RTE expression, data from large human studies are lacking. Here we assessed the expression levels of 795 blood RTE subfamilies in 2467 participants of the population-based Rhineland Study. We found that the expression of more than 98% of RTE subfamilies increased with both chronological and biological age. Moreover, the expression of heterochromatin regulators involved in RTE silencing were negatively related to the expression of 690 RTE subfamilies. Finally, we observed sex differences in 42 RTE subfamilies, with higher expression in men. The genes mapped to sex-related RTEs were enriched in immune response-related pathways. Importantly, we validated our key findings in an independent population-based cohort. Our findings indicate that RTEs and their repressors are markers of aging, and that their dysregulation is linked to inflammation, especially in men.

**Graphical abstract:** 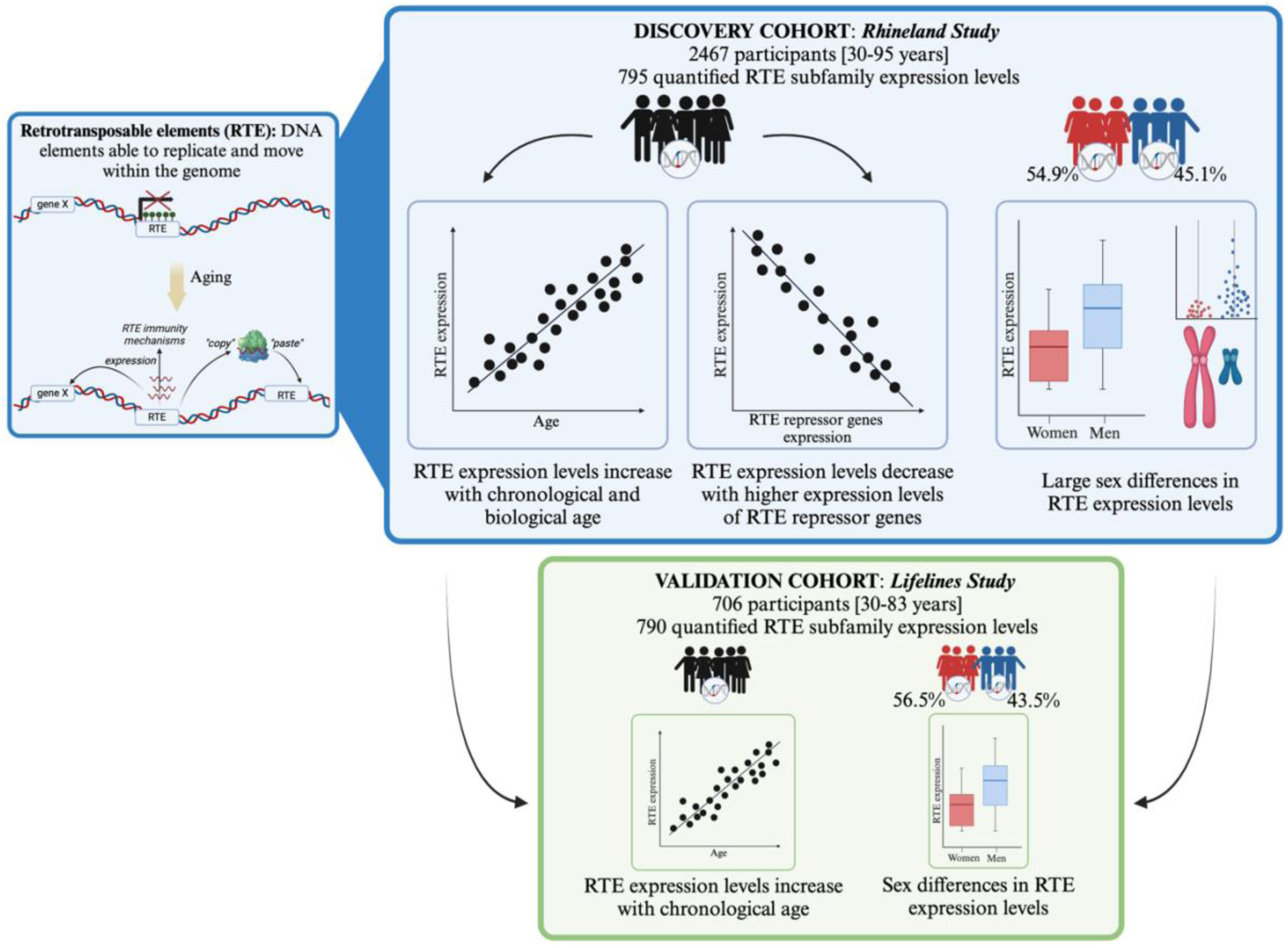

## Introduction

The rapid increase in human life expectancy has spurred interest in understanding of the molecular mechanisms underlying aging and late-life health. Several different approaches have been employed to elucidate the molecular and cellular pathways implicated in aging processes (*1*), focusing mainly on detecting genetic polymorphisms associated with age-related traits (*2*), exploring age-related epigenetic changes (*3*), and defining age-dependent transcriptomic signatures (*4*). In the past few years, thanks to technological advances in sequencing, repetitive segments of DNA previously known as “junk DNA” have been characterized and found to regulate many biological processes (*5, 6*). Indeed, by far the largest proportion of repetitive elements in the human genome is composed of transposable elements (TEs), DNA sequences able to move within the genome (*7*). TEs represent at least 45% of the entire human DNA (*8*), and they are mainly found in the form of retrotransposable elements (RTEs), which use a so-called “copy and paste” mechanism to spread across the genome (*7*). As a result of their long evolutionary history, RTE sequences have accumulated mutations, rendering them widely different. According to sequence homology, transposition mechanisms and structural features, RTEs are classified hierarchically into super-, sub- and separate distinct families (*7*). The quantification of RTE expression was initially limited to the subfamily level. However, more recently further characterization of RTEs at loci level has become feasible, enabling localization and measurement of the expression levels of single RTEs (*9*), although validation of these methods in large genomic datasets is lacking.

Despite the fact that many RTEs in humans have lost their retrotransposition ability due to accrued mutations in the course of evolution, RTEs have been proposed to influence cellular function and tissue homeostasis in the host species via additional mechanisms. Specifically, RTEs could modulate the expression of adjacent genes by acting as enhancers and promoters, while RTEs as RNAs or retrotranscribed DNA could engage the nucleic acid-sensing machineries and trigger inflammatory pathways and antiviral mechanisms (*10–15*). Finally, proteins encoded by RTEs have been linked to senescence and propagation of pathogenic aggregates (*16, 17*). Therefore, to prevent aberrant activation, RTE expression is tightly regulated by several silencing mechanisms, particularly heterochromatin regulation (*18, 19*). An intricate network of factors, including chromatin-modifying enzymes, chromatin-associated proteins, and histone modifications ensure and maintain a proper heterochromatin configuration (*18*). Previous studies, mostly conducted in animal models, reported age-related accumulation of RTEs (*15, 20–23*), induced by decreased efficiency of repressive mechanisms during aging (*24, 25*). Our own work has suggested that derepression of RTEs leads to aging-associated phenotypes, such as myeloid bias and inflammation (*11*). Furthermore, a recent study proposed a causal role of RTEs in aging in *C. elegans* (*26*), while small-scale human studies suggest an association between retrotransposon expression in blood and cellular senescence and inflammation (*27*). Despite substantial progress in the study of aging and aging-related traits in the context of RTEs, previous research has primarily focused on RTEs at the family level (*22, 27*). Limited knowledge exists about the expression of specific RTE subfamilies in aging, and specifically about the localization of individual RTE loci, although the biological role of RTEs is also dependent on their genomic location, having the capability to influence the expression of mapped or neighboring genes (“*cis* effects”) (*28*). Moreover, the amount of RTEs as well as the content of heterochromatin differs, not only across age but also between sexes, mainly due to the presence of large repetitive sequences on the Y-chromosome (*29, 30*). Indeed, aging is a sex-specific process, with sex differences in both lifespan and the occurrence of many diseases (*31*), including inflammatory (*32*), cardiovascular (*33*), metabolic (*34*) and neurodegenerative diseases (*35, 36*). However, scant research is available on potential sex-differences in the expression levels of RTEs in the general population.

Thus, to assess whether dysregulation of RTE expression is a marker of human aging, we evaluated the relation between RTE expression levels and chronological age in a large population-based cohort. Given that deregulation of RTE expression could also be a driver of the aging process and accelerate aging traits (*15, 22, 26, 37*), we also assessed the relation between RTE expression levels and biological age employing “GrimAge”, an estimator of epigenetic aging that is associated with both morbidity and mortality risk (*38*). Indeed, we observed an increase in the expression levels of more than 98% of RTE subfamilies with chronological age. Moreover, we found higher expression of specific RTEs in men compared to women. Importantly, we could replicate the associations of both age and sex with RTE expression levels using data from an independent population-based cohort study. Finally, we were able to demonstrate a negative association between the expression levels of RTE subfamilies and those of chromatin-associated methyltransferase and repressors, key factors for heterochromatin maintenance and RTE silencing (*18*).

## Results

### Discovery study population

A total of 2467 participants were included in our discovery cohort derived from the Rhineland Study (see **Methods** for a detailed description). The mean (standard deviation (SD)) age was 54.95 (14.36) years and 45.1% were men (**Table 1**). We could quantify the expression levels of 795 RTE subfamilies in peripheral blood transcriptome data from participants of the Rhineland Study, which could further be classified into 20 RTE families and 4 RTE superfamilies. SINE superfamily expression represented almost 52% of the whole RTE superfamily expression levels across 2467 participants, followed by LINE, LTR and retroposon superfamilies (**Fig. S1**).

**Table 1.**
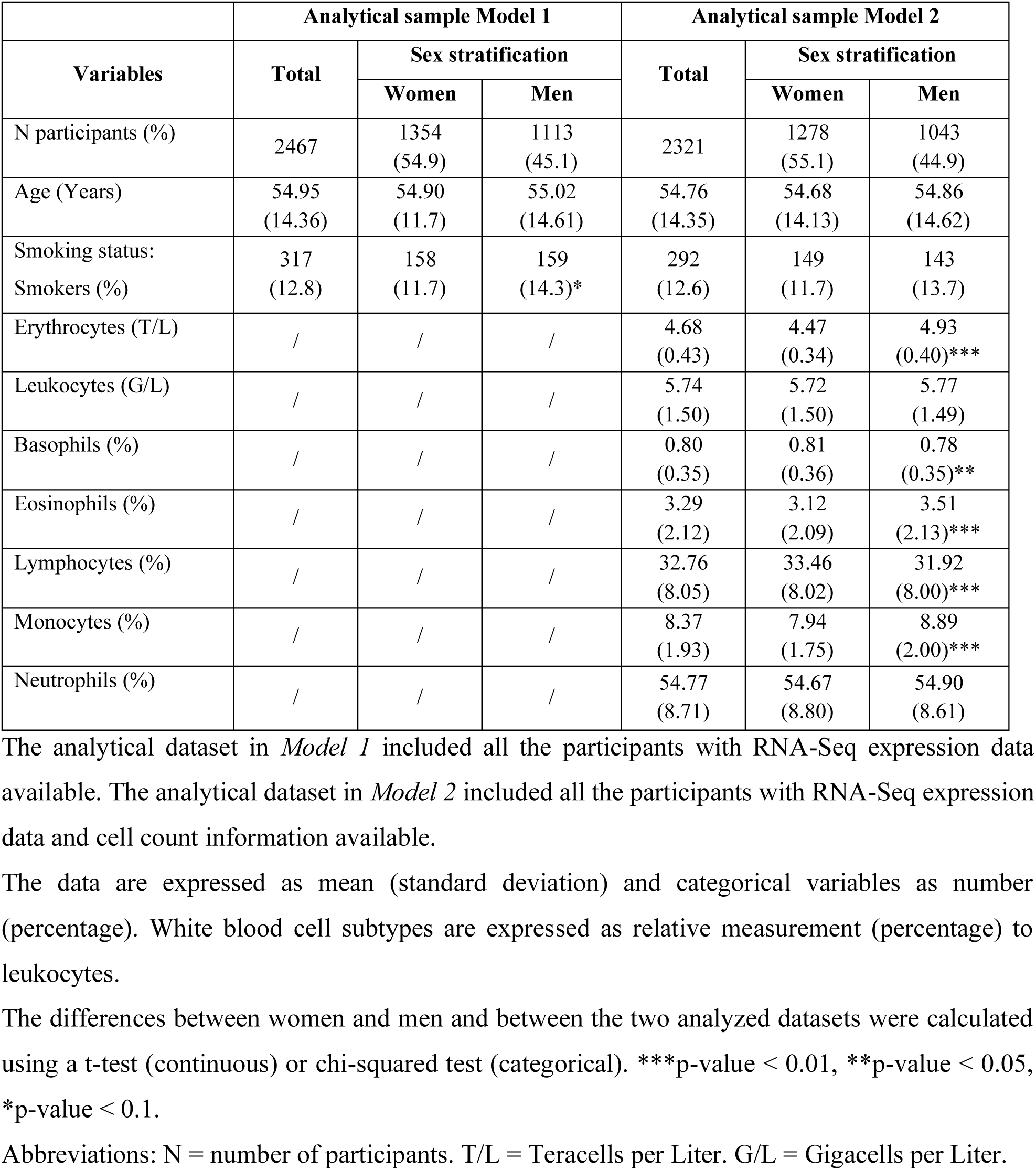
Demographic characteristics of the participants and stratification by sex.

| Variables | Analytical sample Model 1 |  |  | Analytical sample Model 2 |  |  |
| --- | --- | --- | --- | --- | --- | --- |
|  | Total | Sex stratification |  | Total | Sex stratification |  |
|  |  | Women | Men |  | Women | Men |
| N participants (%) | 2467 | 1354<br>(54.9) | 1113<br>(45.1) | 2321 | 1278<br>(55.1) | 1043<br>(44.9) |
| Age (Years) | 54.95<br>(14.36) | 54.90<br>(11.7) | 55.02<br>(14.61) | 54.76<br>(14.35) | 54.68<br>(14.13) | 54.86<br>(14.62) |
| Smoking status:<br>Smokers (%) | 317<br>(12.8) | 158<br>(11.7) | 159<br>(14.3)* | 292<br>(12.6) | 149<br>(11.7) | 143<br>(13.7) |
| Erythrocytes (T/L) | / | / | / | 4.68<br>(0.43) | 4.47<br>(0.34) | 4.93<br>(0.40)*** |
| Leukocytes (G/L) | / | / | / | 5.74<br>(1.50) | 5.72<br>(1.50) | 5.77<br>(1.49) |
| Basophils (%) | / | / | / | 0.80<br>(0.35) | 0.81<br>(0.36) | 0.78<br>(0.35)** |
| Eosinophils (%) | / | / | / | 3.29<br>(2.12) | 3.12<br>(2.09) | 3.51<br>(2.13)*** |
| Lymphocytes (%) | / | / | / | 32.76<br>(8.05) | 33.46<br>(8.02) | 31.92<br>(8.00)*** |
| Monocytes (%) | / | / | / | 8.37<br>(1.93) | 7.94<br>(1.75) | 8.89<br>(2.00)*** |
| Neutrophils (%) | / | / | / | 54.77<br>(8.71) | 54.67<br>(8.80) | 54.90<br>(8.61) |

The analytical dataset in *Model 1* included all the participants with RNA-Seq expression data available. The analytical dataset in *Model 2* included all the participants with RNA-Seq expression data and cell count information available.
The data are expressed as mean (standard deviation) and categorical variables as number (percentage). White blood cell subtypes are expressed as relative measurement (percentage) to leukocytes.
The differences between women and men and between the two analyzed datasets were calculated using a t-test (continuous) or chi-squared test (categorical). \*\*\*p-value < 0.01, \*\*p-value < 0.05, \*p-value < 0.1.
Abbreviations: N = number of participants. T/L = Teracells per Liter. G/L = Gigacells per Liter.

### Changes in RTE expression levels with chronological age

RTE subfamily expression levels increased with chronological age in 97.5% (95% confidence interval (CI): 96.1 to 98.3) of the 795 RTE subfamilies (one proportion Z-test p-value < 2.2e-16) (*Model 1*). However, only two subfamilies (i.e., MER54B and LTR43-int) retained statistical significance (false discovery rate (FDR) < 0.05) after multiple comparisons adjustment. The same positive trend was also observed after adjusting for blood cell composition (*Model 2*), indicating higher expression levels in 99.5% (95% CI: 98.7 to 99.8) of RTE subfamilies with older chronological age (one proportion Z-test p-value < 2.2e-16). Similar to *Model 1*, only MER54B was significantly positively associated with chronological age after FDR-adjustment (**Fig. 1A**, **Data S1**, **Fig. S2A**). The per locus analysis, conducted on the 64 MER54B loci, revealed that four of these loci were the main drivers of the association with chronological age. The expression levels of three of these loci increased with age and they were located in intronic regions or overlapped with splicing sites of the mapped genes. Specifically, two loci were located in the beta-defensin gene cluster on chromosome 8, while the others were positioned within the *NBEA* and *RAB27B* genes (**Fig. 1B**, **Data S2**, **Fig. S2B**). Further evaluation of the mapped and *cis*-genes revealed that the expression levels of three age-related loci were positively associated with the expression levels of the corresponding genes. Specifically, the expression levels of the two loci on chromosome 8 were positively associated with those of the *cis*-gene *DEFA3,* while the expression levels of the RTE locus within *NBEA* gene showed a strong association with those of the gene itself, suggesting co-expression of these two transcripts (**Data S3**).

**Fig. 1.**
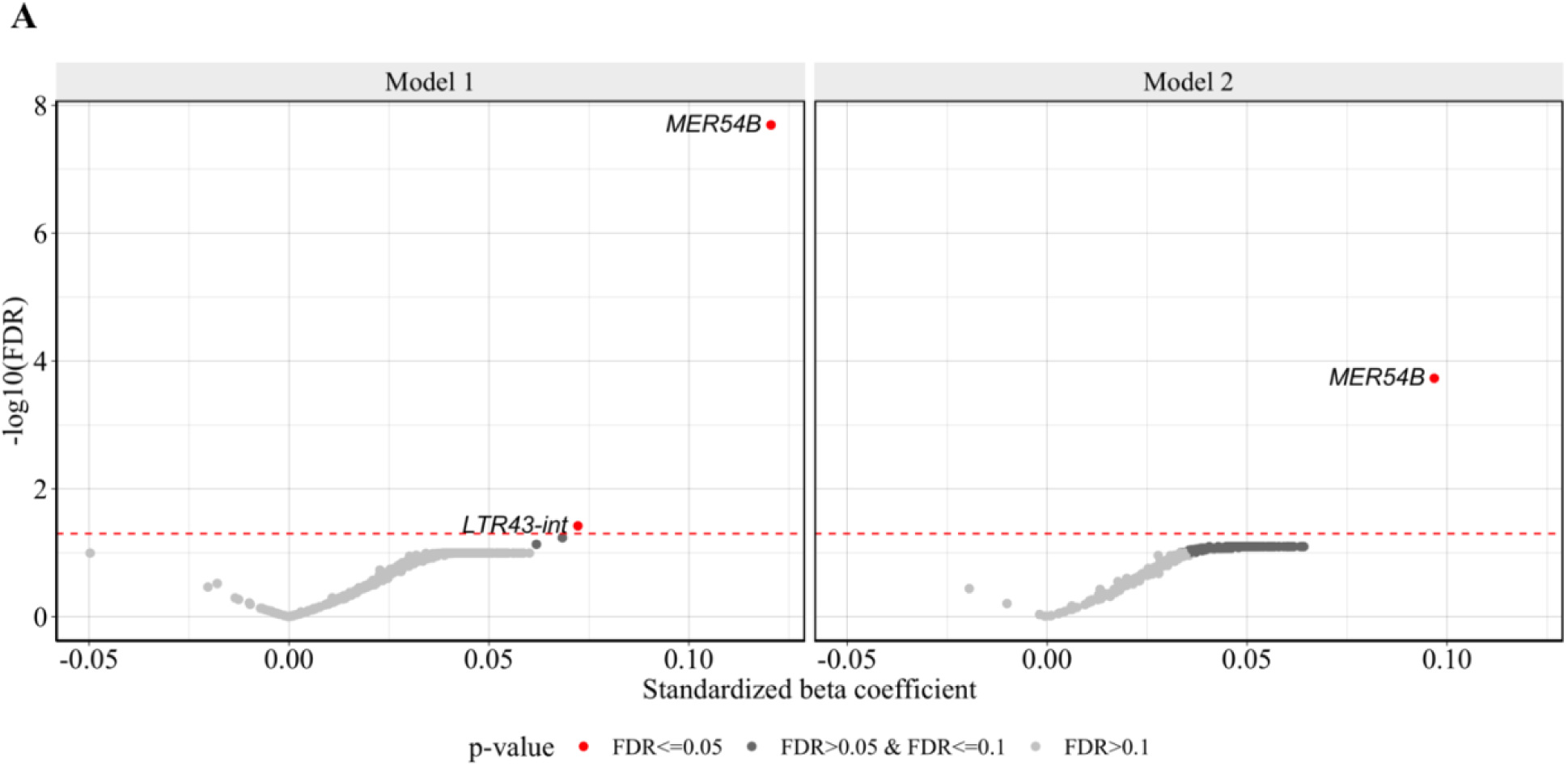
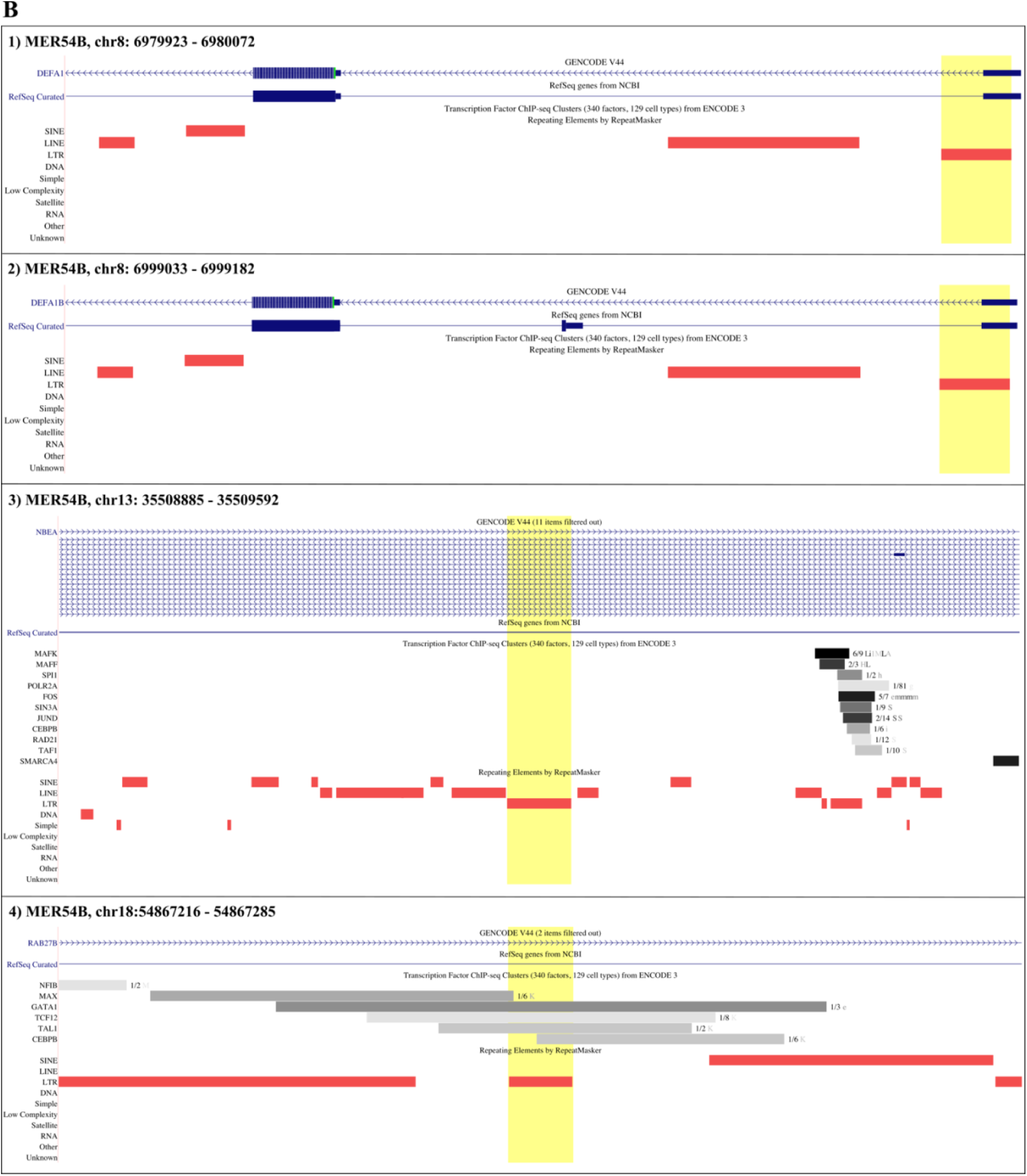
Association of RTE subfamilies and loci with chronological age. **(A)** Volcano plot showing the change in the standardized expression of RTE subfamilies per one standard deviation in chronological age. Each dot represents a RTE subfamily. The red horizontal line indicates the threshold for statistical significance, set at FDR<0.05. *Model 1*: RTE subfamily_i_ ∼ intercept + age + sex + smoking + batch. *Model 2*: Additionally adjusted for blood cell composition. **(B)** Visualization of the four MER54B age-associated RTE loci, highlighted by the yellow shading, in the human genome reference GRCh38/hg38 through UCSC Genome Browser. Mapped genes (in blue) and transcription factor binding sites (in grey) are also indicated, if present.

The sensitivity analysis, conducted on three additional RTE subfamilies that were most strongly associated with chronological age (i.e., CR1.L3A-Croc, AluYk11 and SVA_C), showed that two CR1.L3A-Croc loci were positively associated with chronological age (**Data S2**). Moreover, we observed that one of these loci, located in chromosome 20, was also positively associated with the corresponding mapped gene, *TSHZ2* (**Fig. S3**, **Data S3**).

Given that the evolutionary youngest human-specific RTE subfamilies (including L1HS and L1PA2) are likely to be functionally most relevant in terms of transposition capacity (*39*), we also performed a separate locus-specific analysis for these RTEs. This analysis revealed that chronological age was significantly associated with one locus (out of 226) for L1HS and nine loci (out of 602) for L1PA2 (**Fig. S4**, **Data S2**). Most of these loci were located in intronic regions and were positively associated with the corresponding mapped or *cis*-genes, including *CAMK4*, *ATM* and *IFI16* (**Data S3**).

### Changes in RTE expression levels with biological age

The relation between RTE subfamilies and biological age mirrored the one observed for chronological age. The expression levels of 99.3% (95% CI: 98.5 to 99.7) of RTE subfamilies increased with higher biological age (one proportion Z-test p-value < 2.2e-16) (*Model 1*). Specifically, 650 RTE subfamilies were associated with biological age at FDR < 0.05. A similar trend was also observed after adjustment for blood cell composition, with 98.0% (95% CI: 96.8 to 98.7) of RTE subfamilies showing increased expression levels with higher biological age (one proportion Z-test p-value < 2.2e-16). However, after correcting for multiple comparisons to minimize false positive rates, none of the associations between separate RTE subfamilies and biological age remained statistically significant (**Fig. 2**, **Data S4**).

**Fig. 2:**
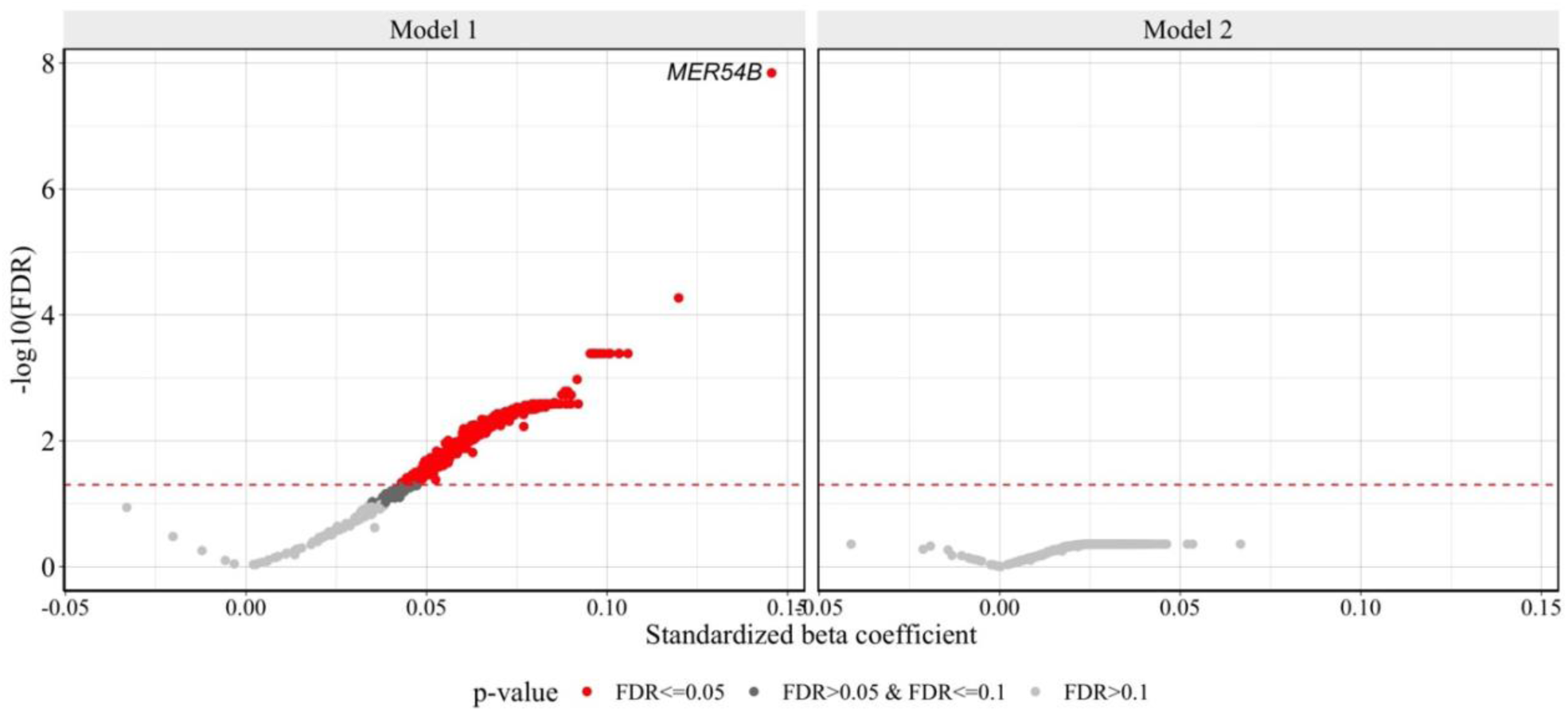
Associations of RTE subfamilies with biological age. Volcano plot showing the change in the standardized expression of RTE subfamilies per one standard deviation in biological age. Each dot represents a RTE subfamily. The red horizontal line indicates the threshold for statistical significance, set at FDR<0.05. *Model 1*: AgeAccelGrim ∼ intercept + RTE subfamily_i_ + sex + smoking + batch. *Model 2*: Additionally adjusted for blood cell composition.

### Sex differences in RTE expression levels

Sex was significantly associated with the expression levels of 55 RTE subfamilies (FDR < 0.05, *Model 1*). After adjustment for blood cell counts, 42 RTE subfamilies remained significantly associated with sex, with higher expression levels in men compared to women (**Fig. 3A**, **Data S5**). With the exception of a single RTE subfamily (L1MDb), the entirety of the sex-related RTE subfamilies belonged to the LTR superfamily, mainly comprised of human endogenous retroviruses (HERVs). The per locus level analysis revealed that these associations with sex were driven by 247 RTE loci. Moreover, it indicated that the higher expression of RTE subfamilies in men was mainly due to the location of 168 of the RTE loci on the Y-chromosome, confined to a window of about 18 million base pairs (**Fig. 3, B and C**, **Data S6**). The genomic annotation of these sex-related loci revealed that they were located mainly in intronic and intergenic regions (**Fig. S5**). By investigating the relation of the RTE loci’s expression with the corresponding mapped and/or *cis*-genes, we found that the majority of the associations were in the same direction, with an increase in RTE loci expression levels corresponding to an increase in gene expression levels. This finding suggests that RTE loci, which are located in intronic regions, are co-expressed with the genes they are mapped to, whereas those located in intergenic regions likely function as regulators of the proxy genes (**Data S7**). Further functional analysis of genes associated with sex-related RTEs revealed significant enrichment for immune responses (e.g., “positive regulation of interleukin-1 beta production” and “cellular response to interferon-beta”) (**Fig. S6**). Sex did not modify the associations of expression levels of RTE subfamilies with chronological age or biological age.

**Fig. 3.**
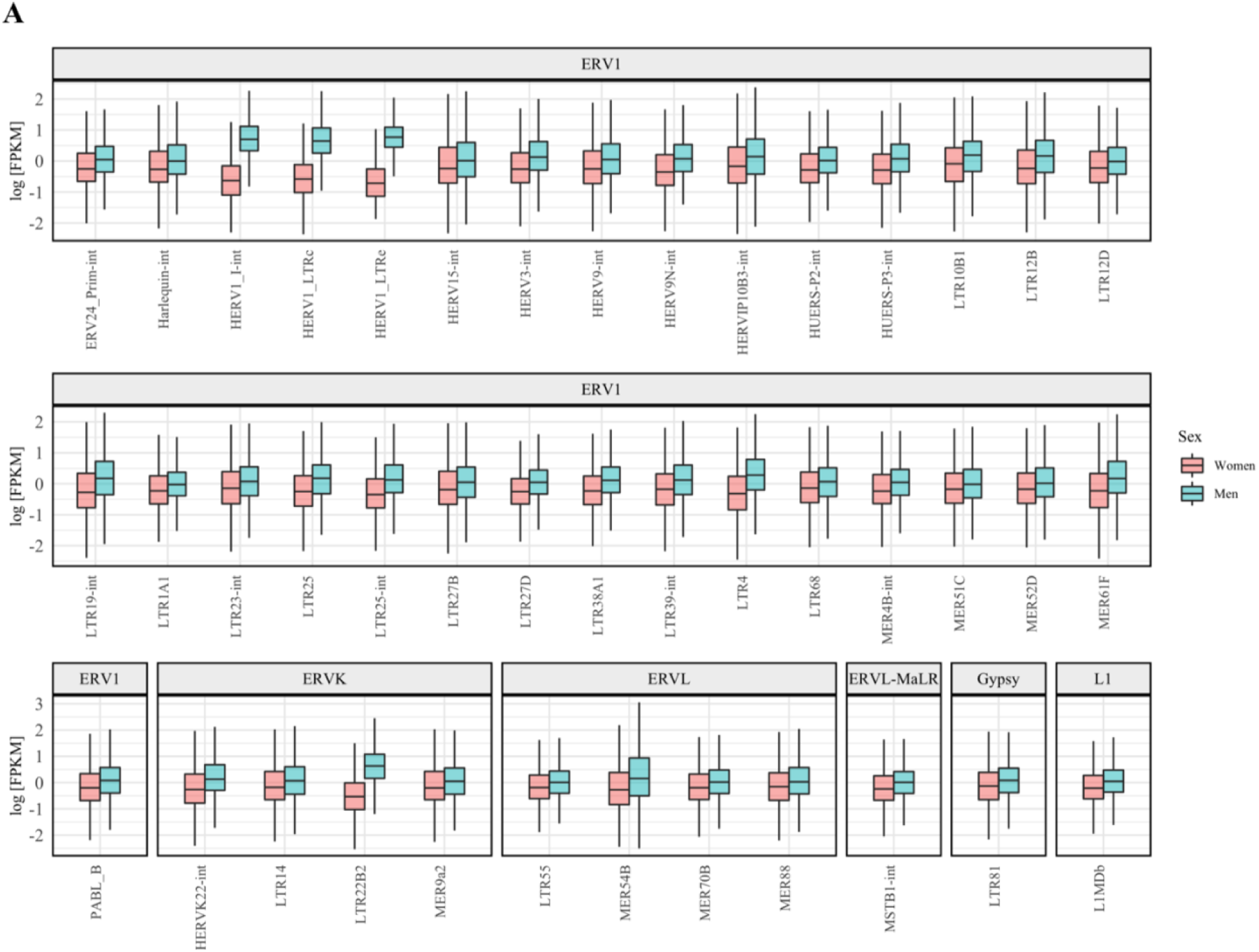
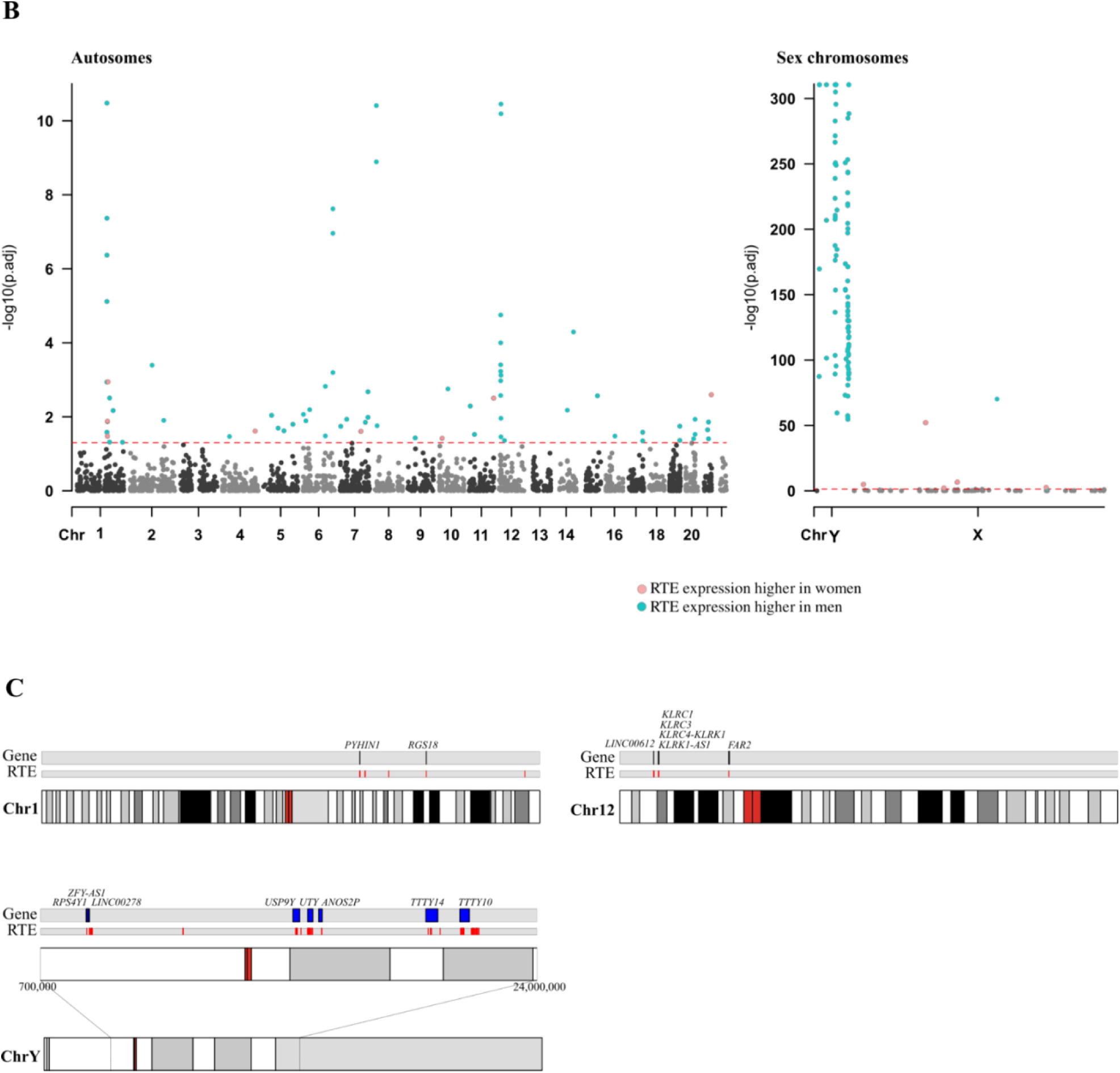
Significant association of RTE subfamilies and loci with sex. **(A**) Box plots showing the different expression levels between men and women for the 42 RTE subfamilies for which we observed a significant sex-difference in the RTE expression levels. The RTE subfamilies are grouped based on the family they belong to. **(B)** Manhattan plot representing RTE loci belonging to the 42 RTE subfamilies associated with sex. The red line indicates the threshold for statistical significance, set at FDR<0.05. **(C)** Ideograms of chromosomes 1, 12 and Y showing the locations of sex-related loci on the respective chromosomes and the mapped genes, when present.

### Sensitivity analysis

The associations of the RTE scores, computed at family, superfamily and overall levels with chronological age, biological age and sex mirrored the ones identified at the subfamily level, confirming the positive association of RTE expression levels with both chronological and biological age, as well as the higher expression levels of RTEs in men compared to women. (**Fig. 4**, **Data S8-S10**).

**Fig. 4:**
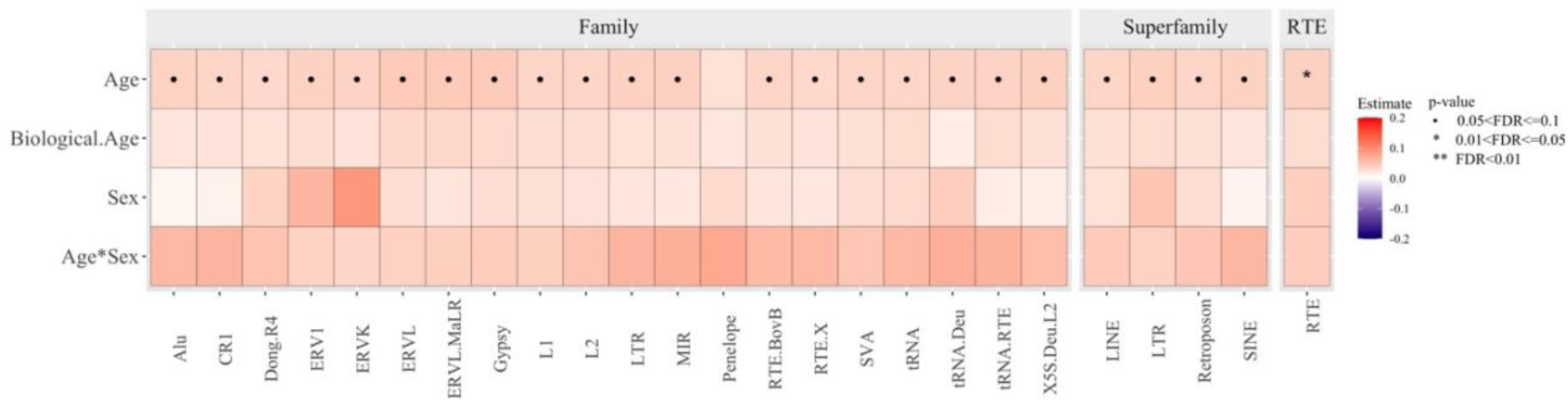
Association of RTE families, superfamilies and overall RTE expression levels with age, sex and biological age. Heatmap representing the strength and the directionality of the associations between RTE family, superfamily and overall RTE expression levels with chronological age, sex, biological age and age-sex interaction.

### Association of RTE repressor genes with RTE subfamilies

The analysis of the effect of heterochromatin regulators on RTE subfamilies revealed that the RTE repressor score (calculated by combining the expression levels of *DAXX*, *TRIM28*, *SETDB1b* and *DNMT1* (*11, 40*)) is negatively associated with the expression levels of age- and sex-related RTE subfamilies (**Fig. 5, Data S11**). Interestingly, this relation was consistent across most RTE subfamilies. Specifically, a lower RTE repressor score corresponded to higher expression levels of 690 RTE subfamilies at FDR < 0.05 (*Model 2*) (**Fig. S7**). Moreover, we found that the RTE repressor score tended to be lower in older than younger participants (ß estimate: −0.11, p-value = 0.099). However, no association with biological age and no sex differences in the RTE repressor scores were observed.

**Fig. 5:**
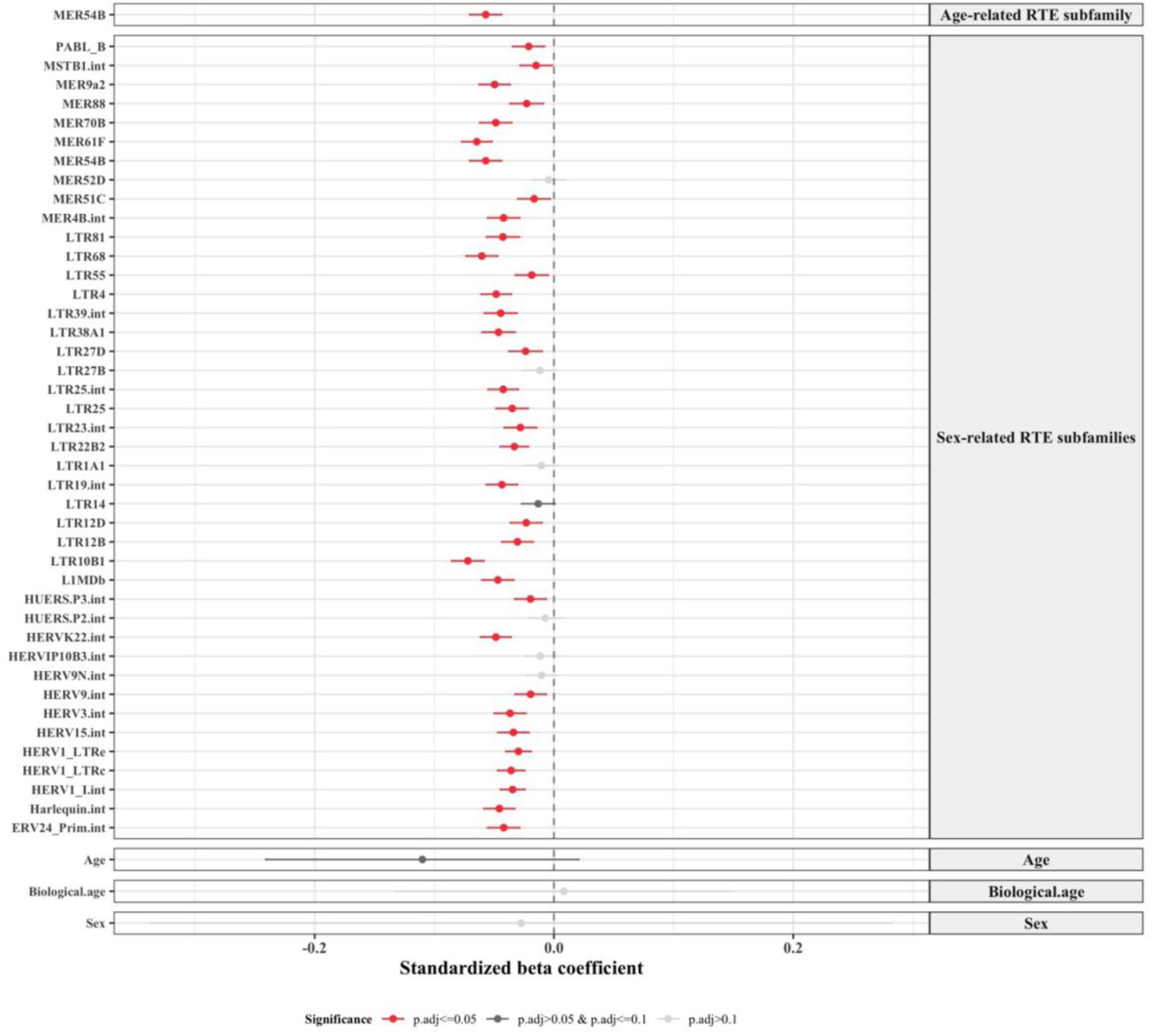
Relation between RTE repressor score and age- and sex-related RTE subfamilies, chronological age and sex. The forest plot shows the associations of RTE repressor score with the age-related RTE subfamily, MER54B, the 42 sex-related RTE subfamilies, chronological age and sex. The standardized beta coefficients for RTE subfamilies were obtained through the following linear regression model: RTE subfamily_i_ ∼ intercept + RTE repressor score + age + sex + smoking + batch + blood cell composition. The standardized beta coefficients for age and sex were obtained through the following linear regression model: RTE repressor score ∼ intercept + age + sex + smoking + batch + blood cell composition. The standardized beta coefficients for biological age and sex were obtained through the following linear regression model: RTE repressor score ∼ intercept + biological age + sex + smoking + batch + blood cell composition.

### Validation of findings in an independent cohort

For external validation of our results, we used data from a total of 706 participants from the population-based Lifelines Study for whom RTE expression levels, age, sex, smoking status and blood cell counts were available (see **Methods** for a detailed description). The mean age of the included participants was 47.7 years (SD: 11.1), with 43.5% male. The expression levels of 790 RTE subfamilies could be quantified in the Lifelines participants.

The age analysis revealed that chronological age was significantly associated with eight RTE subfamilies at p-value < 0.05 (*Model 1*) (**Data S12**). The RTE subfamily MER54B, identified as the top hit associated with chronological age in the Rhineland Study, was validated in the Lifelines cohort (ß estimate: 0.112, p-value = 0.005). Additionally, we observed a positive trend between chronological age and the expression levels of RTE subfamilies, consistent with the findings from the Rhineland Study. Specifically, expression levels increased with chronological age in 93.2% (95% CI: 91.2 to 94.7) of the 790 RTE subfamilies (one proportion Z-test p-value < 2.2e-16). After adjusting for blood cell concentrations (*Model 2*), the results remained consistent, with chronological age still showing a positive association with the expression levels of eight RTE subfamilies at p-value < 0.05, including the MER54B subfamily (**Table 2**). Additionally, similar to *Model 1*, 92.3% (95% CI: 90.2 to 93.9) of the 790 RTE subfamilies exhibited increased expression levels with chronological age (one proportion Z-test p-value < 2.2e-16), mirroring the findings of the Rhineland Study (**Data S12**).

**Table 2.**
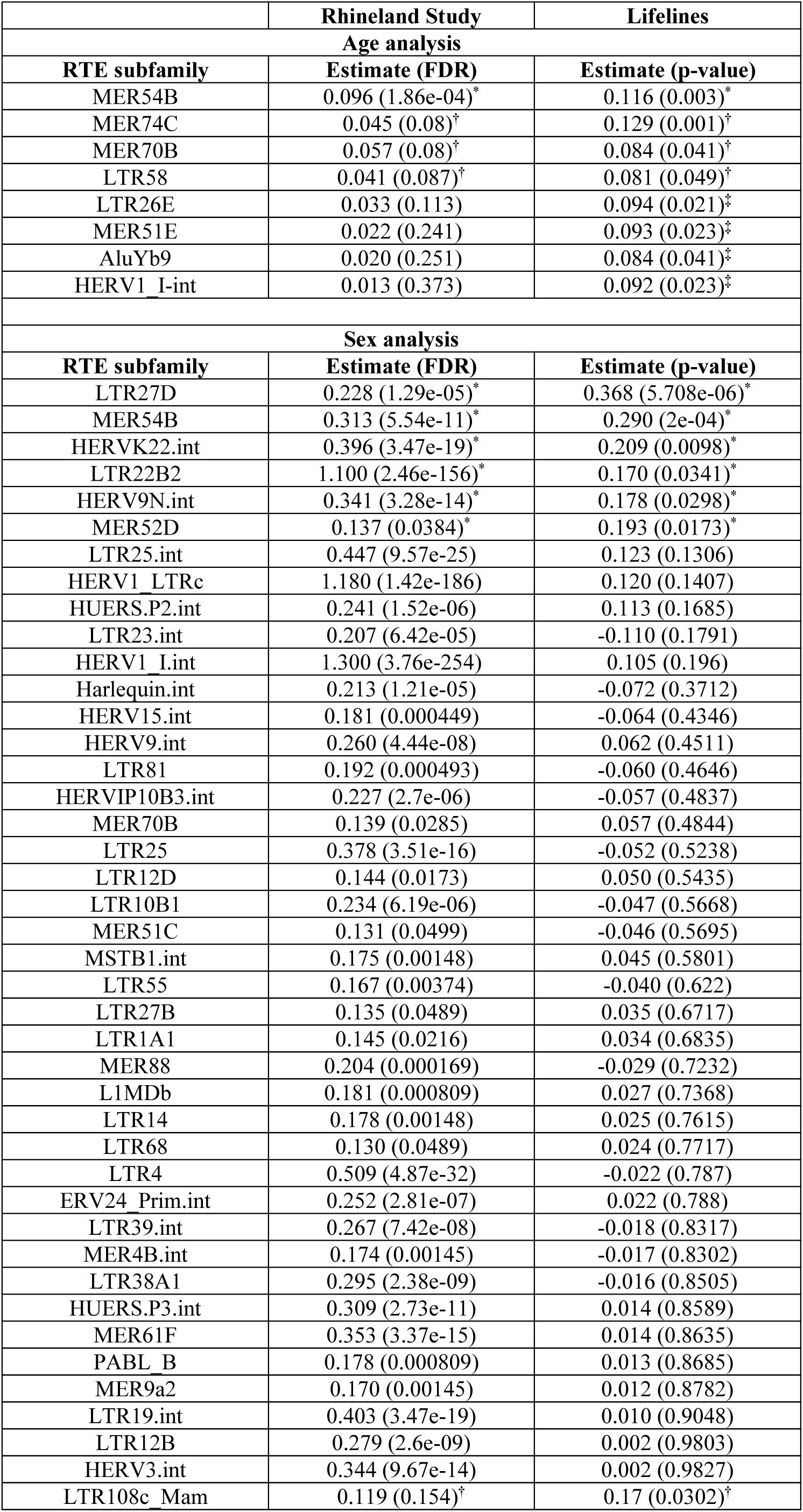
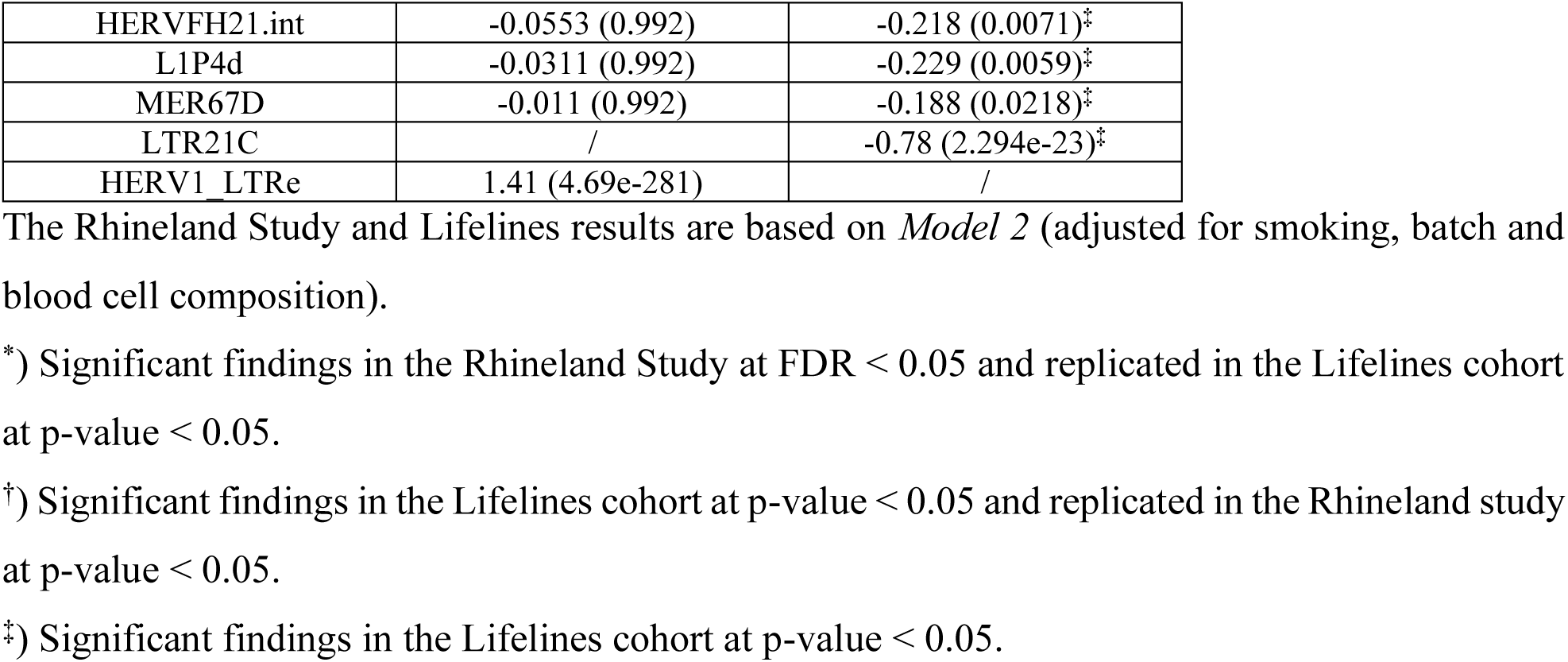
Validation of the Rhineland Study findings in the Lifelines cohort.

The sex analysis in the validation cohort also indicated that sex was significantly associated with the expression levels of 13 RTE subfamilies in *Model 1* and 11 RTE subfamilies in *Model 2* at p-value < 0.05. Among these, the sex-related associations observed in the Rhineland Study were replicated for 7 and 6 of these RTE subfamilies in *Model 1* and *2*, respectively (**Table 2, Data S13**).

## Discussion

To our knowledge, this is the first study reporting a comprehensive overview of genome-wide RTE subfamily expression levels in blood across age and sex in the general population, with further detailed RTE locus-specific characterization. We found that for more than 98% of RTE subfamilies expression levels increased with both higher chronological and biological age. The chronological age-related findings were replicated in an independent cohort, where also more than 92% of RTE subfamilies exhibited higher expression levels with increasing age. Moreover, we found higher levels of RTE expression in men compared to women, mostly due to the expression of LTR elements on the Y-chromosome, which was also largely replicated in the independent Lifelines cohort. Interestingly, the expression of sex-related RTE loci was primarily associated with the expression of genes involved in immune response pathways. In addition, our results revealed a relation between elevated expression levels of heterochromatin regulator genes and reduced expression levels of RTE subfamilies, indicating that transposable elements are tightly regulated by epigenetic silencing mechanisms.

We observed higher genome-wide expression levels of RTE subfamilies with older chronological age. This association was also apparent when summing up the RTE subfamily expression levels per (super)family, indicating that RTE levels and activity increase with age. The increased expression levels of different RTE families with advancing age has also been reported in animal models (*20, 23*) and small-scale human studies (*22*). Despite reporting an association between RTE expression and biological age-related events, a recent study did not find a link between chronological age and RTE expression (*27*), which is likely due to the study’s relatively small sample size and use of microarray expression data, precluding comprehensive analyses at RTE subfamily level. Increased RTE expression with age is thought to be primarily linked to heterochromatin loss (*24*), due to the reduction of histone proteins (*41*), decreased DNA methylation (*42*), different histone variants and modifications (*43, 44*), as well as dysregulation of histone chaperons, such as *DAXX* (*45*). Among all the RTE subfamilies, the expression levels of MER54B, classified in the ERVL (LTR) family, showed the strongest association with chronological age. Human endogenous retroviruses (HERVs) are transposable elements originating from the insertion of exogenous retroviruses in the germ line. Most of them are nonfunctional with respect to transposition due to accumulated mutations during evolution (*46*). Nonetheless, HERVs have been associated with several pathological processes, although the exact underlying mechanisms are still unknown.

It has also been suggested that HERVs might act as transcriptional regulatory elements for neighboring genes (*47*). In support of this hypothesis, our per locus analysis identified two age-related MER54B loci, located within the cluster of defensin genes, whose expression was positively associated with that of the *cis*-gene *DEFA3.* Defensin genes encode for peptides specialized in host innate immune and infection responses (*48*), and they have been reported as potential inflammatory biomarkers of frailty (*49*). The co-expression of age-related RTE loci and genes involved in inflammatory pathways further supports the contribution of these elements to the aging process itself (*16*). Moreover, we identified another MER54B age-related locus within the intronic region of *RAB27B*, a gene responsible for the production of endogenous exosomes, key regulators of the immune response (*50*). Indeed, through multimodal experiments, previous studies demonstrated that increased HERVK expression as well as the accumulation of HERVK-Env proteins and HERVK retrovirus-like particles triggers the innate immune response and promotes the intercellular spreading of proteopathic seeds (*16, 17*). The fourth MER54B age-related locus was located in the intronic region of the *NBEA* gene, which encodes the autism-related protein neurobeachin involved in the regulation of synaptic protein trafficking (*51*). Additionally, several *de novo NBEA* genetic variants have been related to early childhood epilepsy (*52*). Interestingly, ERVs have also been shown to increase genomic instability and play a critical role in the pathogenesis of autism-spectrum disorders (*53*). However, to what extent similar mechanisms may contribute to age-associated traits, especially cognitive decline, remains to be established. Nonetheless, the strong positive association between the expression levels of this particular RTE locus and *NBEA* indicates that co-localization of RTE loci within or in the vicinity of genes results in their co-transcription (*54*).

Interestingly, the expression levels of the youngest human-specific subfamilies, belonging to the long interspersed nuclear element-1 (L1) family, increased with chronological age. Specifically, L1HS, also known as L1PA1, is the youngest human-specific subfamily (*8, 39, 55*) and has retained its active retrotransposing ability (*56*). We found that the age-related L1HS locus is located within the *CAMK4* gene. The CaMK4 protein is a multifunctional serine/threonine kinase, involved in various aspects of the immune response. It plays a critical role in regulating the production of cytokines, including interleukin-17 and has been implicated in the pathogenesis of autoimmune diseases such as systemic lupus erythematosus and psoriasis (*57, 58*). L1PA2 is the evolutionary second youngest human-specific subfamily. The corresponding age-related loci were interspersed across the genome, either located within the intronic regions or adjacent to various genes, including *ATM* and *IFI16.* The ATM protein, a serine/threonine protein kinase, plays a key role in regulating the cellular response to DNA damage by activating multiple DNA repair pathways (*59*). Dysfunction of ATM can lead to genomic instability and cellular senescence (*60*). Meanwhile, the IFI16 protein, an interferon gamma inducible protein, is involved in the immune response by detecting viral DNA and activating the interferon signaling pathway (*61, 62*). Although we did not directly quantify RTE transposition activity, the higher expression levels of L1HS and L1PA2 subfamilies with chronological age may reflect their increased transposition activity during aging.

The increase of RTE subfamily expression levels with advancing chronological age was also mirrored in the analysis with biological age as the outcome. We observed that an acceleration of biological age corresponded to higher expression values of almost all RTE subfamilies. This result supports the hypothesis that RTE expression levels could serve as transcriptomic markers of aging (*22*). In fact, our findings indicate that not only the activity of RTE subfamilies increases with the actual number of years a person has lived, but also seems to reflect the aging process. This result is in line with the findings reported by LaRocca et al., where the expression of repetitive elements was found to be a good predictor of age (*22*).

We not only found differences in the expression of RTE subfamilies across age, but also between sexes. Expression levels of 42 RTE subfamilies were higher in men compared to women. A relatively large number of sex-related RTE subfamilies belonged to the ERV1 family. It has been suggested that HERVs’ retroviral DNA/RNA directly activates inflammatory responses(*14*), as also suggested by several enriched biological processes in our pathway analysis (e.g., “positive regulation of interleukin-1 beta production” and “cellular response to interferon-beta”). The higher expression of RTE subfamilies in men than women was mainly due to the presence of 168 expressed loci on the Y-chromosome. This result is consistent with the evidence that the Y-chromosome is a hot spot for RTEs, as reported in animal and human studies (*29, 30*). However, it is still unknown why the Y-chromosome contains a relatively large number of repeat elements. One plausible explanation could be the presence of non-recombining genomic regions on the Y-chromosome (*63*).

Besides RTE loci located on the Y-chromosome, we also found several RTE loci located on autosomes, which might also contribute to differences in the biology of men and women. These autosomal sex-related loci were mainly located in the promoter and intronic regions of chromosomes 1 and 12. Although the expression of most RTE loci was higher in men compared to women, the LTR25 locus, located in an intergenic region of chromosome 1, was one of the few loci with a higher expression in women compared to men. The expression levels of this RTE locus were positively associated with, among others, those of *cis*-genes belonging to the cluster Fc Gamma receptor genes (*FCGR2B, FCGR2C, FCGR3B, FCGR3A*). This suggests that the LTR25 locus acts as a regulator of the neighboring genes, controlling the expression of Fc Gamma receptor genes, which is in line with the fact that women have higher antibody responses compared to men (*32*). Given that women are generally more susceptible to inflammatory diseases (*32*), the small number of RTE loci exhibiting relatively higher expression levels in females is intriguing, and might reflect the presence of sex-specific RTE silencing mechanisms that remain to be elucidated. A non-mutually exclusive hypothesis would be that the relatively higher levels of LTRs in men could play a physiological role in limiting the immune response in males, potentially contributing to sex-based differences in immune regulation.

Interestingly, for both age- and sex-related RTE subfamilies, we observed a decrease in their expression levels with higher expression levels of heterochromatin regulator genes. This result aligns with the knowledge that the activity of RTEs is tightly regulated to prevent aberrant expression (*64*). The maintenance of heterochromatin integrity is the main mechanism that ensures silencing of transposable elements. Several molecules are part of the intricate machinery that is responsible for keeping chromatin highly compact, including DNA and histones (e.g., methyltransferase enzymes (e.g., *DNMT*s, *SETDB1*) and the *KRAB-ZPF-TRIM28* gene complex (*64–66*)). Previous studies have already shown that the depletion of any of these factors might result in an increase of RTE activity (*67, 68*), mainly attributed to the perturbation of chromatin structure. Our findings reinforce the importance of this phenomenon by demonstrating that within the general population individuals with relatively higher levels of heterochromatin regulator genes had lower expression levels of RTE subfamilies.

This study has both strengths and limitations. We investigated the effects of age and sex on blood RTE subfamily expression levels in a large population-based study, including a large number of healthy individuals across a wide age range (30-95 years old), with further validation of the key findings in an independent cohort. In addition, candidate RTE subfamilies were further characterized by comprehensively evaluating all their comprising RTE loci across the genome. Moreover, we explored their potential biological role by relating them to the expression levels of nearby genes. Finally, incorporating heterochromatin regulator genes into the analysis yielded important clues to the potential biological mechanisms underlying the silencing process of RTEs. However, due to the cross-sectional nature of our study, we were unable to assess the temporal directionality of the effects, though our findings are highly consistent with those from previous experimental studies. Additional longitudinal and experimental studies are needed to further explore the biological role of RTEs as modulators of aging and age-related diseases.

In conclusion, we found that blood RTE expression levels increased with both chronological and biological age and were higher in men compared to women. Additionally, increased expression of RTEs was associated with decreased expression of heterochromatin regulator genes and, in men, with increased expression levels of neighboring genes involved in immune response. Collectively, our findings indicate that RTEs are intimately associated with the aging process, and that their dysregulation may be linked to inflammation, especially in men.

## Materials and Methods

### Discovery study population

For the discovery phase of our study, we used data from participants of the Rhineland Study, an on-going population-based cohort study in Bonn, Germany. Invitations are sent to individuals from two geographically defined areas in Bonn who are eligible for inclusion, i.e., aged 30 years and above and have sufficient command of the German language to provide informed consent. The study was approved by the ethics committee of the University of Bonn, Medical Faculty. The Rhineland Study is carried out in accordance with the recommendations of the International Conference on Harmonization (ICH) Good Clinical Practice (GCP) standards (ICH-GCP). Written informed consent was obtained from all participants in accordance with the Declaration of Helsinki. We based our study on data from the first 3000 consecutively enrolled participants of the Rhineland Study. The current analysis included 2467 participants for whom RNA sequencing (RNA-Seq) data was available after quality control.

### Measurements

#### Whole blood RNA isolation and sequencing

We extracted total RNA from peripheral blood. Blood was collected in PAXgene Blood RNA tubes (PreAnalytix/Qiagen) and processed according to manufacturer’s guidelines. PAXgene tubes were thawed and incubated at room temperature to increase RNA yields. Total RNA was isolated according to manufacturer’s instructions using PAXgene Blood miRNA Kit and following the automated purification protocol (PreAnalytix/Qiagen). RNA integrity and quantity was evaluated using the tapestation RNA assay on a Tapestation 4200 instrument (both from Agilent). We used 750 ng total RNA to generate NGS libraries using the TruSeq stranded total RNA kit (Illumina) following manufacturer’s instructions with Ribo-Zero Globin reduction. We checked library size distribution via Tapestation using D1000 on a Tapestation4200 instrument (Agilent) and quantified the libraries via Qubit HS dsDNA assay (Invitrogen). We clustered the libraries at 250 pM final clustering concentration on a NovaSeq6000 instrument using S2 v1 chemistry (Illumina) in XP mode and sequenced paired-end 2*50 cycles before demultiplexing using bcl2fastq2 v2.20. Quality control of the sequencing was evaluated through FastQC (v0.11.9).

#### Retrotransposable element subfamily and loci profiling

To quantify the expression of retrotransposable elements (RTEs) in RNA-Seq data, we used the SQuIRE pipeline (Software for Quantifying Interspersed Repeat Elements)(*9*). SQuiRE quantifies expression of retrotransposable elements at both the subfamily and locus level. The reads were aligned to the human reference genome hg38 from UCSC and RTE annotation was based on RepeatMasker (*69*). The quantification step uses an iterative method to quantify unique and multi-mapping reads. SQUIRE outputs both read counts and FPKM (fragments per kilobase transcript per million reads) for RTE subfamily as well as for RTE loci. RTE subfamilies which were expressed in at least 5% of the participants with an average of mapped reads equal or bigger than 15 were considered for further analyses, in order to improve sensitivity since low expressed RTEs might reflect noise in the quantification phase. We log-ransformed FPKM counts to conform the data to a normal distribution.

#### Gene expression profiling

To quantify the expression of genes in RNA-Seq data, the sequencing reads were first aligned to the human reference genome GRCh38.p13 provided by Ensembl using STAR v2.7.1. The count matrix was generated with STAR –quantMode GeneCounts using the human gene annotation version GRCh38.101. Genes with an overall mean expression greater than 15 reads and expressed in at least 5% of the participants were used for the following analyses. Finally, in order to normalize for library size and to log-transform the raw data, we applied the varianceStabilizingTransformation function from DESeq2 v1.30.1 R package.

#### Estimation of RTE family and superfamily expression scores and overall RTE expression

RTE family and superfamily expression levels were estimated by first standardizing the expression levels of RTE subfamilies and then summing the Z-scores of the RTE subfamilies belonging to the same family/superfamily. Moreover, we obtained the overall RTE expression levels by summing the Z-scores of the 795 RTE subfamilies.

#### DNA methylation quantification and biological age estimation

Genomic DNA was extracted from buffy coat fractions of anti-coagulated blood samples using Chemagic DNA buffy coat kit (PerkinElmer, Germany) with Chemagic Magnatic Separation Module 1 and Chemagic Prime 8 Automated Workstation, and was subsequently bisulfite converted using the EZ-96DNA Methylation-Lightning^TM^MagPrep from Zymo according to the manufacturer’s instructions. DNA methylation levels were measured on Illumina iScan using Illumina’s Human MethylationEPIC BeadChip. Sample-level and probe-level quality control was performed using the ‘minfi’ package in R (version 3.5.0). GrimAge was calculated based on the algorithm developed by Lu et al., using 1030 CpG sites. Biological age was defined as the residual (in years) that results from regressing GrimAge on chronological age (*38*).

#### Demographic and biochemical variables

Data on age, sex and smoking status were based on self-reports. Smoking status was defined as a dichotomous variable (e.g., current smoker and not-current smoker). Missing values for smoking status were imputed using cotinine metabolite levels, measured through the Metabolon HD4 platform (*70*). Differential blood cell counts (e.g., erythrocytes, leukocytes, basophils, eosinophils, lymphocytes, monocytes, neutrophils) were performed at the Central Laboratory of the University Hospital in Bonn using EDTA-whole blood samples on a hematological analyzer Sysmex XN9000. For the analysis of biological age, defined as the described below, blood cell type proportions, estimated from methylation data, were used as covariates (*71*).

### Statistical analysis

#### Association of chronological age and sex with RTE subfamily expression levels

We performed multivariable regression analyses to assess the association of chronological age and sex with the expression levels of each RTE subfamily. We also tested possible age and sex interaction effects on RTE subfamily expression levels. As covariates, we included smoking status and batch (*Model 1*). To account for blood cell composition, we further adjusted the model for white and red blood cell counts, and the relative fractions of basophils, eosinophils, lymphocytes, monocytes, and neutrophils (*Model 2*). All the continuous variables were standardized in order to enable comparisons among variables with different units. To account for multiple comparisons, the results from the regression models were corrected using the false discovering rate (FDR) method. The level for statistical significance was set at FDR<0.05.

#### RTE subfamily expression levels and biological age

Multivariable regression was also used to test the association of expression levels of RTE subfamilies with biological age. First, we regressed out the batch effect of methylation from biological age. Next, we used the biological age residuals as the outcome to investigate its relation with RTE subfamilies. As previously, *Model 1* was adjusted for sex, smoking status and batch of RNA-Seq data. To further account for changes of blood composition with age and/or sex, we additionally adjusted for white and red blood cell counts and the relative fractions of CD8^+^ T cells, CD4^+^ T cells, natural killer cells, B cells, monocytes, and granulocytes estimated from methylation data (*Model 2*).

#### Association of RTE locus expression levels with chronological age and sex

To identify which RTE loci were driving the effects of age and/or sex, for RTE subfamilies that were significantly associated with age or sex (at FDR<0.05), we additionally assessed the associations of age and sex with RTE expression at the level of each single locus. To this end, we employed a zero-inflated Gaussian mixed-model in order to account for excessive zeros in the expression data of RTE loci. Specifically, the expression at each RTE locus was used as the outcome, and age and sex were included as independent variables. All models were adjusted for smoking status, white and red blood cell counts, as well as relative fractions of different white blood cell subtypes, including batch as a random effect. The level for statistical significance was set at FDR<0.05. As a further sensitivity analysis, we also performed single locus analyses for those RTE subfamilies that exhibited the largest effect sizes for the association with chronological age, selecting one from each superfamily (i.e., short interspersed nuclear element (SINE), long interspersed nuclear element (LINE), long terminal repeat (LTR) and retroposon).

#### Relation of RTE locus expression levels with mapped/CIS genes

Using multivariable regression, we also investigated the association of candidate RTE loci (independent variables) with mapped and *cis*-genes (outcome variables), while adjusting for age, sex, batch, smoking status, white and red blood cell counts and the relative fractions of white blood cell subtypes. *Cis-*genes were defined as those genes lying within a 100 Kb window (upstream or downstream) of a candidate RTE locus and a p-value of < 0.05 was considered statistically significant for this targeted approach. Pathway enrichment analysis was conducted on genes which were significantly associated with the corresponding RTE locus. The pathway enrichment analysis was performed using the “clusterProfiler” R package (v. 3.18.1), querying Gene Ontology: Biological Processes (GO:BP). Redundant terms collected from Gene Ontology were grouped using the rrvgo R Bioconductor package (v. 1.2.0) with default parameters.

#### Sensitivity analysis

To investigate whether the changes in RTE expression levels across different subfamilies were additive with regard to chronological age, biological age and sex, we performed additional regression analyses using *Model 1* and *Model 2* as described previously, but now using the summary scores of RTE expression as outcomes (i.e., RTE family Z-score, RTE superfamily Z-score and overall RTE Z-score as defined above).

#### Exploring the associations of “RTE repressor” genes with RTE subfamilies

To evaluate the association of age- and sex-dependent RTE subfamilies with genes involved in the maintenance of heterochromatin structure, we selected four key methyltransferase and co-repressor genes whose expression levels were available in the Rhineland Study transcriptomics dataset (i.e., *DAXX*, *TRIM28*, *SETDB1b* and *DNMT1*)(*11*). Specifically, we created a summary score for these four “RTE repressor” genes by regressing out batch effects from the expression levels of each gene followed by summing up of the Z-standardized residuals of the four genes. A multivariable linear regression analysis was then performed to explore the association between the RTE repressor score (main independent variable) and each age- and sex-related RTE subfamily (dependent variable), while adjusting for age, sex and smoking status (*Model 1*), and additionally for white and red blood cell counts, and the relative fractions of basophils, eosinophils, lymphocytes, monocytes, and neutrophils (*Model 2*). The significance level was set at FDR<0.05. All the statistical analyses were performed in R 4.0.3.

### External validation

We used data from a subset of the Lifelines Study, an independent population-based cohort study, to validate our key results regarding the associations of both chronological age and sex with the expression levels of different RTE subfamilies. The Lifelines Study is a large population-based cohort study in the north of the Netherlands, which collects deep pheno- and genotypic data (*72*). To ensure comparability with the participants of the Rhineland Study, we only included participants aged 30 years and above. Of these, 706 had both RNA-seq and the required covariate data available and were included in our validation study.

In Lifelines, RNA was isolated from whole blood, collected in PAXgene tubes using the PAXgene Blood miRNA Kit (Qiagen, California, USA). Illumina’s Hiseq2000 instrument was used for sequencing with paired-end 2×50 bp reads. Detailed information about the RNA isolation and sequencing process is described in the LifeLines DEEP manuscript (*73*). To obtain the RTE subfamily counts in the Lifelines dataset, we applied the SQuIRE pipeline to the RNA-Seq data, using the same normalization and filtering steps as described for the Rhineland Study data. Smoking status was collected through questionnaires, while the concentrations of blood cells (i.e., leukocytes, mononuclear cells and neutrophilic granulocytes) were assessed in plasma extracted from fresh blood samples at the laboratory center of the University Medical Center Groningen.

We assessed the association of chronological age and sex with the expression levels of RTE subfamilies through multivariable regression analyses, adjusting for smoking status (*Model 1*), and additionally for leukocytes, neutrophiles and monocytes concentrations (*Model 2*). All the continuous variables were standardized. For validation of the associations that survived FDR-correction in the discovery cohort, the level of statistical significance was set at p-value < 0.05 in the replication cohort.

## Data Availability

The Rhineland Study's dataset is not publicly available because of data protection regulations. Access to data can be provided to scientists in accordance with the Rhineland Study's Data Use and Access Policy. Requests for further information or to access the Rhineland Study's dataset should be directed to. Lifelines data may be obtained from a third party and are not publicly available. Researchers can apply to use the Lifelines data used in this study. More information about how to request Lifelines data and the conditions of use can be found on the Lifelines study website (https://www.lifelines-biobank.com/researchers/working-with-us).

## Acknowledgments

We would like to thank all participants of the Rhineland Study and the study personnel involved in the extensive data collection and all members of PRECISE for performing RNA-sequencing. We would like to also thank the Lifelines Cohort study and its contributing research centers for providing the data, as well as to all the study participants.

## Funding

NAA is partly supported by a European Research Council Starting Grant (Number: 101041677). The Rhineland Study is funded by the German Center for Neurodegenerative Diseases (DZNE). This work was further supported by the Deutsche Forschungsgemeinschaft (DFG, German Research Foundation) under Germany’s Excellence Strategy (EXC2151-390873048) and SFB 1454 (Project-ID 432325352); through the Federal Ministry of Education and Research grant (FKZ: 01KX2230; “PreBeDem – Mit Prävention und Behandlung gegen Demenz”), and the Helmholtz Association under the 2023 Innovation Pool. DL is partly supported by a grant from the Alzheimer’s Association (24AARFD-1192360). PS is currently being supported by the following funding bodies: the DZNE and the Ministry of Culture and Science of the State of Northrhine Westphalia (CANcer TARgeting, CANTAR programme as part of Netzwerke 2021) and the German Cancer Aid Program THUNDER. Work in part related to the topic of this manuscript was supported by the ERC (Consolidator Award H3.3Cancer, number 616744), the Wilhelm Sander Stiftung, the Worldwide Cancer Research Fund (WCRF), the DFG under Germany’s Excellence Strategy (Grant no. EXC2151–390873048; Excellence Cluster ImmunoSensation2), the Helmholtz-Gemeinschaft Aging and Metabolic Programming (AMPro) Consortium, and along with other funding bodies. PS is recipient of an Honorary Professorship at UCL (2023-2027). PS thanks current and former members of his group for discussions, in particular Dr Jenny Russ and Dr Julia Gerber. MDB is supported by the excellence cluster ImmunoSensation2 (EXC 2151, #390873048); the DFG via IRTG2168 (#272482170), SFB1454 (#432325352); the EU-funded project NEUROCOV receiving funding from the RIA HORIZON Research and Innovation under GA No. 101057775; the Else-Kröner-Fresenius Foundation (2018_A158).

The Lifelines initiative has been supported by funding from the Dutch Ministry of Health, Welfare and Sport, the Dutch Ministry of Economic Affairs, the University Medical Center Groningen (UMCG), Groningen University, and the provinces in the north of the Netherlands (Drenthe, Friesland, Groningen).

## Author contributions

Conceptualization: VT, HL, DL, PS, MMBB, NAA. Methodology: VT, HL, DL, MDB, PS, MMBB, NAA. Formal analysis: VT. Investigation: VT. Data acquisition: MDB, MMBB, NAA. Data curation: VT, DL. Validation: PS, MMBB, NAA. Supervision: PS, MMBB, NAA. Project administration: MMBB, NAA. Funding acquisition: MMBB. Writing – original draft: VT, HL, DL, PS, MMBB, NAA. Writing – review and editing: MDB.

## Competing interests

Authors declare that they have no competing interests.

## Notes

### Competing Interest Statement

The authors have declared no competing interest.

### Author Declarations

The study was approved by the ethics committee of the University of Bonn, Medical Faculty. The Rhineland Study is carried out in accordance with the recommendations of the International Conference on Harmonization (ICH) Good Clinical Practice (GCP) standards (ICH-GCP). Written informed consent was obtained from all participants in accordance with the Declaration of Helsinki.

