## Supplemental Figures for "Age and sex effects on blood retrotransposable element expression levels: Findings from the population-based Rhineland Study"

Fig. S1. Overview of the average RTE superfamily and family expression levels in the Rhineland Study.

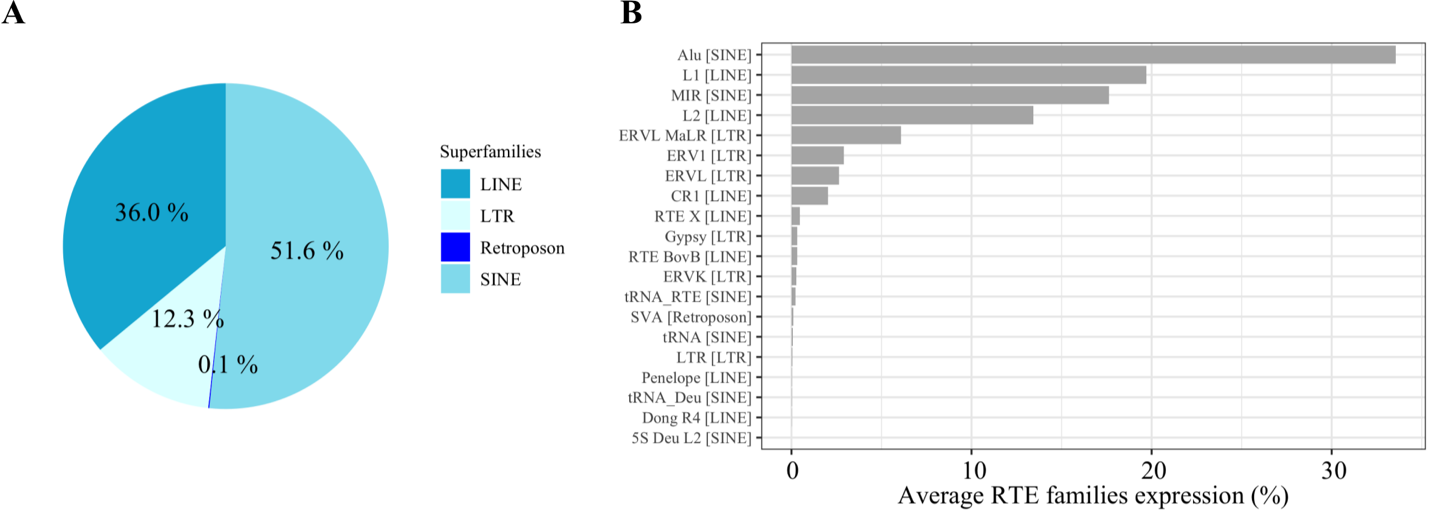

The average expression levels for RTE superfamilies and families were calculated as the sum of the average expression of the corresponding RTE subfamily across the entire population. **(A)** The pie-plot shows the average expression level (reported as percentage value) of each RTE superfamily in the Rhineland Study. **(B)** The bar-plot represents the expression level (reported as percentage value) of each RTE family in the Rhineland Study. The family names on the y-axis are followed by the corresponding superfamily names in square brackets.

**Fig. S2. Overview of the key RTE subfamilies and loci analyzed.**

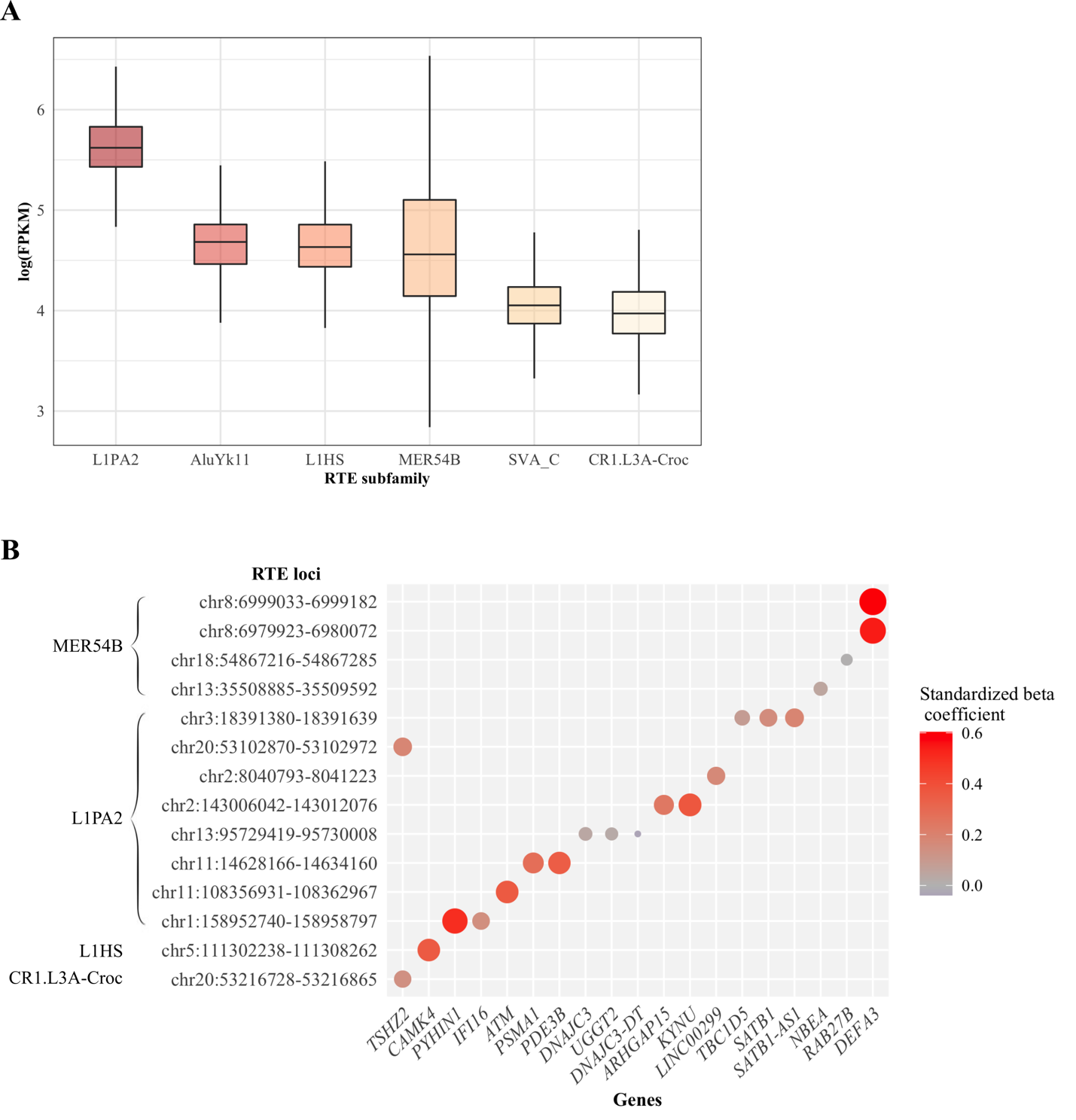

**(A)** Box plot showing the expression levels of selected RTE subfamilies analyzed in this study. **(B)** Heatmap representing the magnitude and directionality of the associations between key RTE loci and the corresponding mapped/*cis*-genes.

Abbreviations: FPKM = fragments per kilobase transcript per million reads

**Fig. S3.** **Sensitivity analysis on three additional RTE subfamilies exhibiting the largest relative effect sizes for the association with chronological age.**
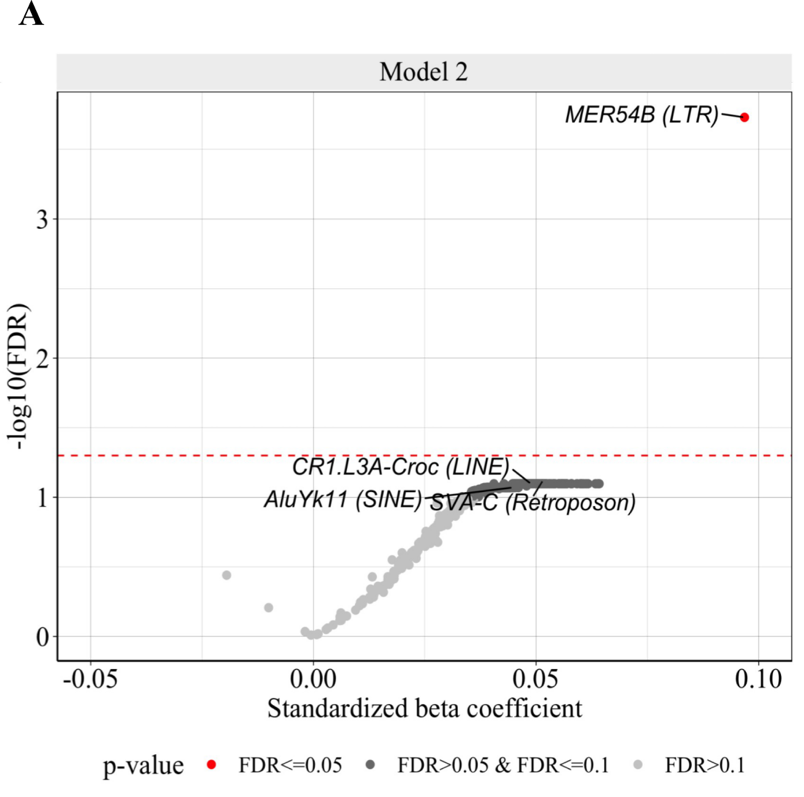

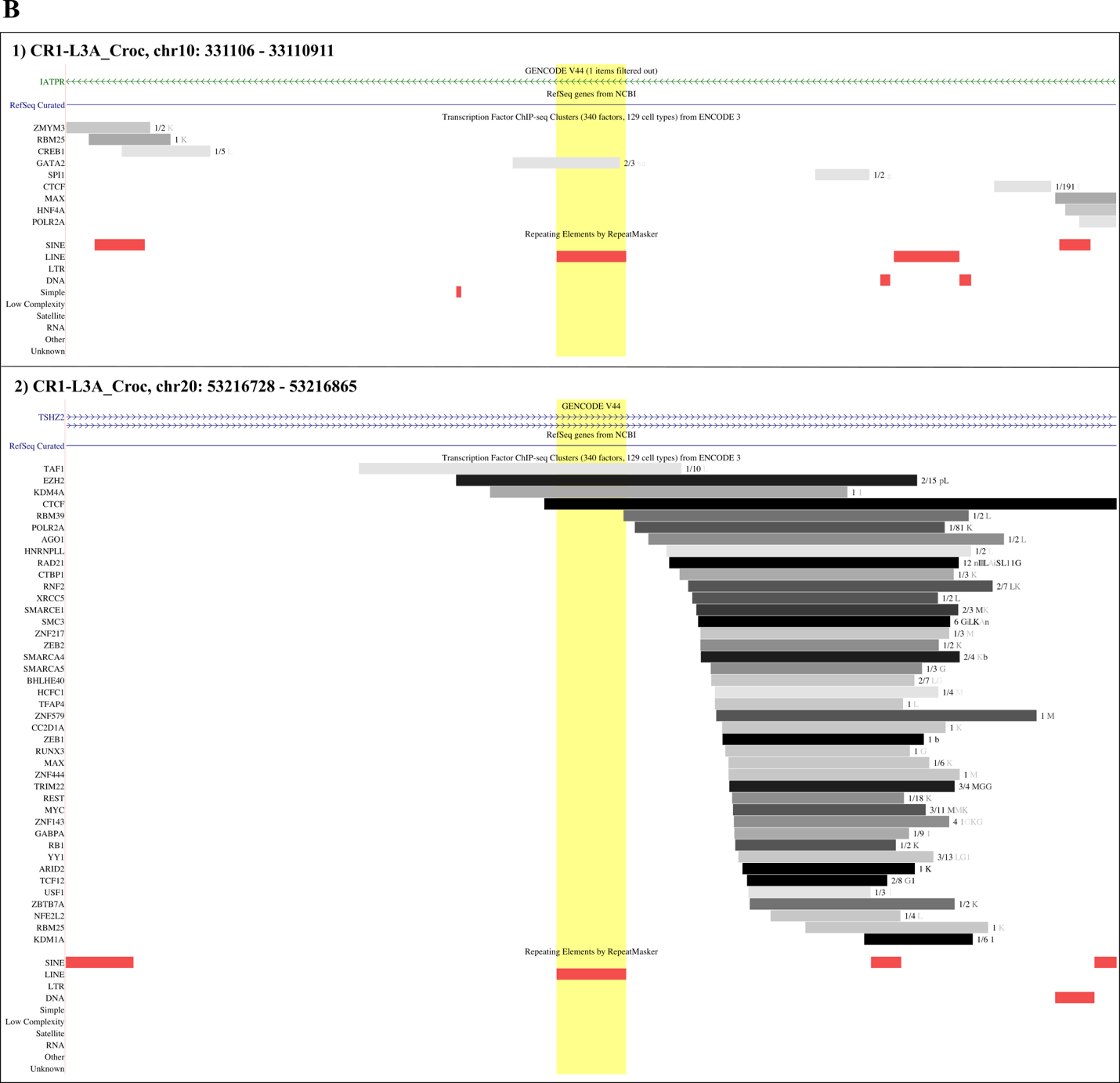

**(A)** Volcano plot showing the change in the standardized expression of RTE subfamilies per one standard deviation in chronological age. The three RTE subfamilies, selected for sensitivity analysis, are labeled in the plot, along with the MER54B subfamily. *Model 2*: RTE subfamily_i ~_ intercept + age + sex + smoking + batch + blood cell composition. **(B)** Visualization of the two CR1.L3A-Croc age-associated RTE loci, highlighted by the yellow shading, in the human genome reference GRCh38/hg38 through UCSC Genome Browser. Mapped genes (in blue) and transcription factor binding sites (in grey) are also indicated if present.

**Fig. S4. Characterization of age-related loci for L1HS and L1PA2, the youngest human-specific RTE subfamilies.**

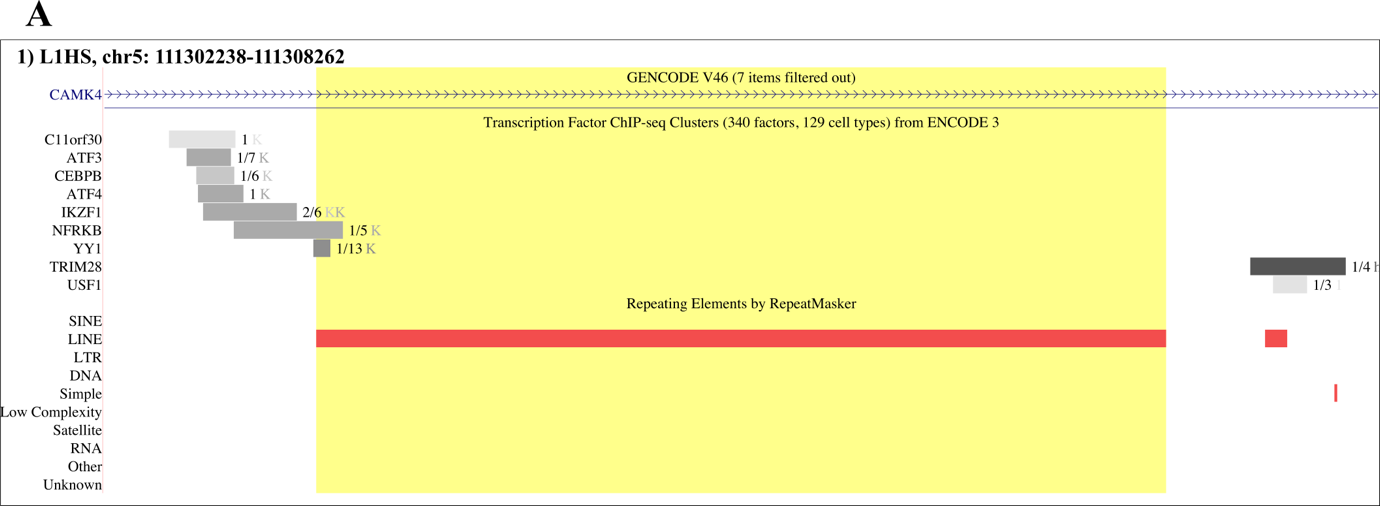

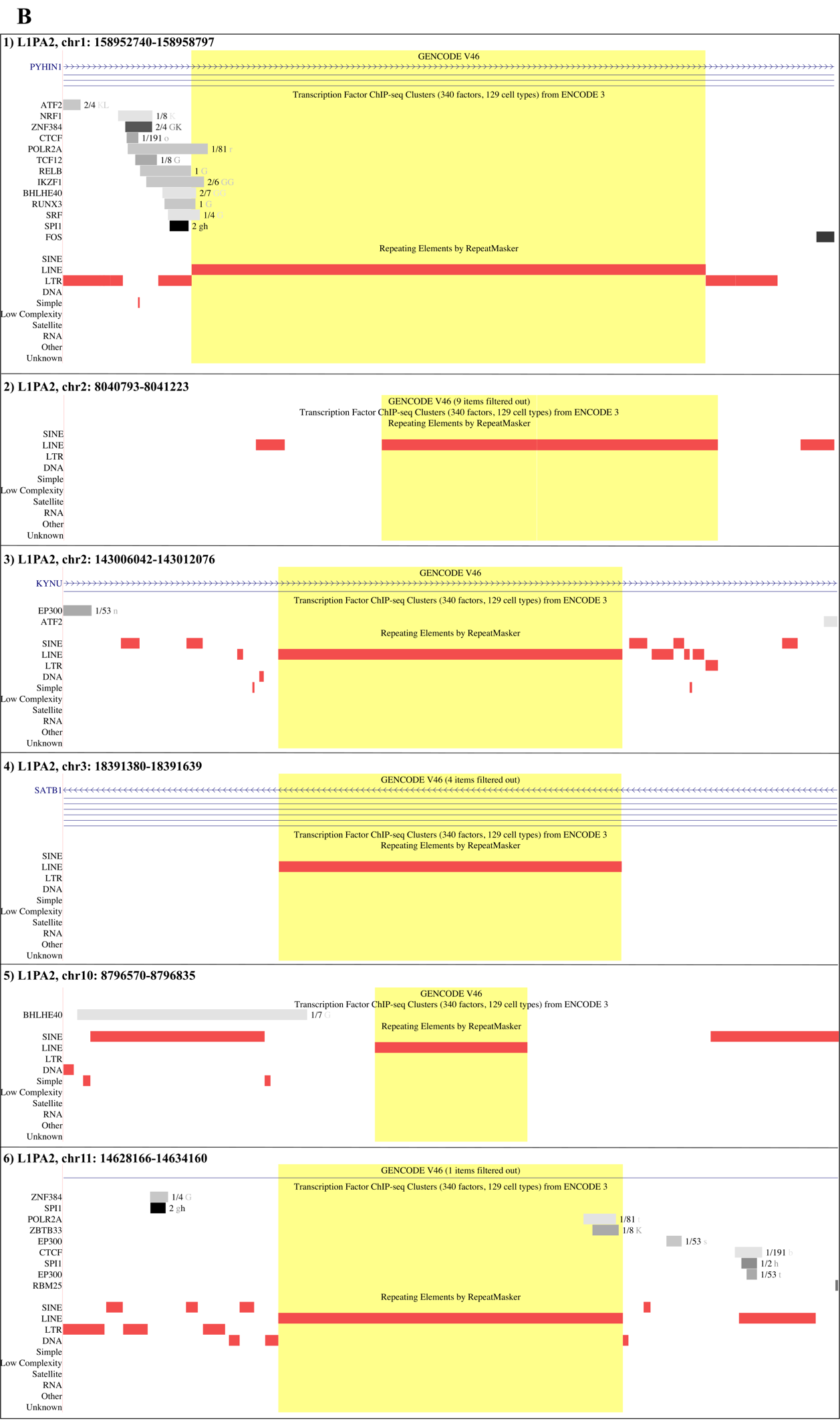

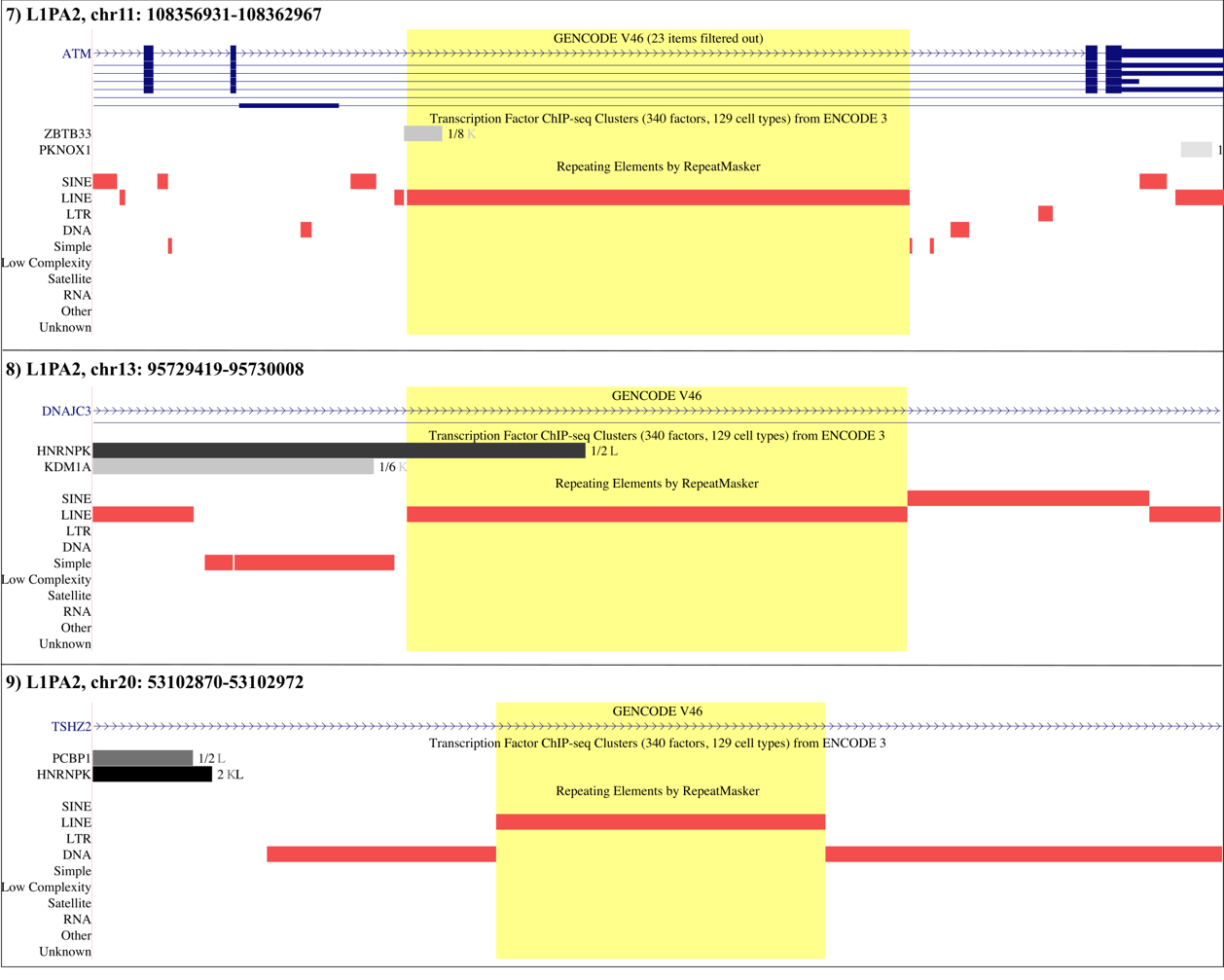

Visualization of **(A)** L1HS and **(B)** L1PA2 age-associated RTE loci, highlighted by the yellow shading, in the human genome reference GRCh38/hg38 through UCSC Genome Browser. Mapped genes (in blue) and transcription factor binding sites (in grey) are also indicated if present.

**Fig. S5. Characterization of sex-associated RTE loci.**
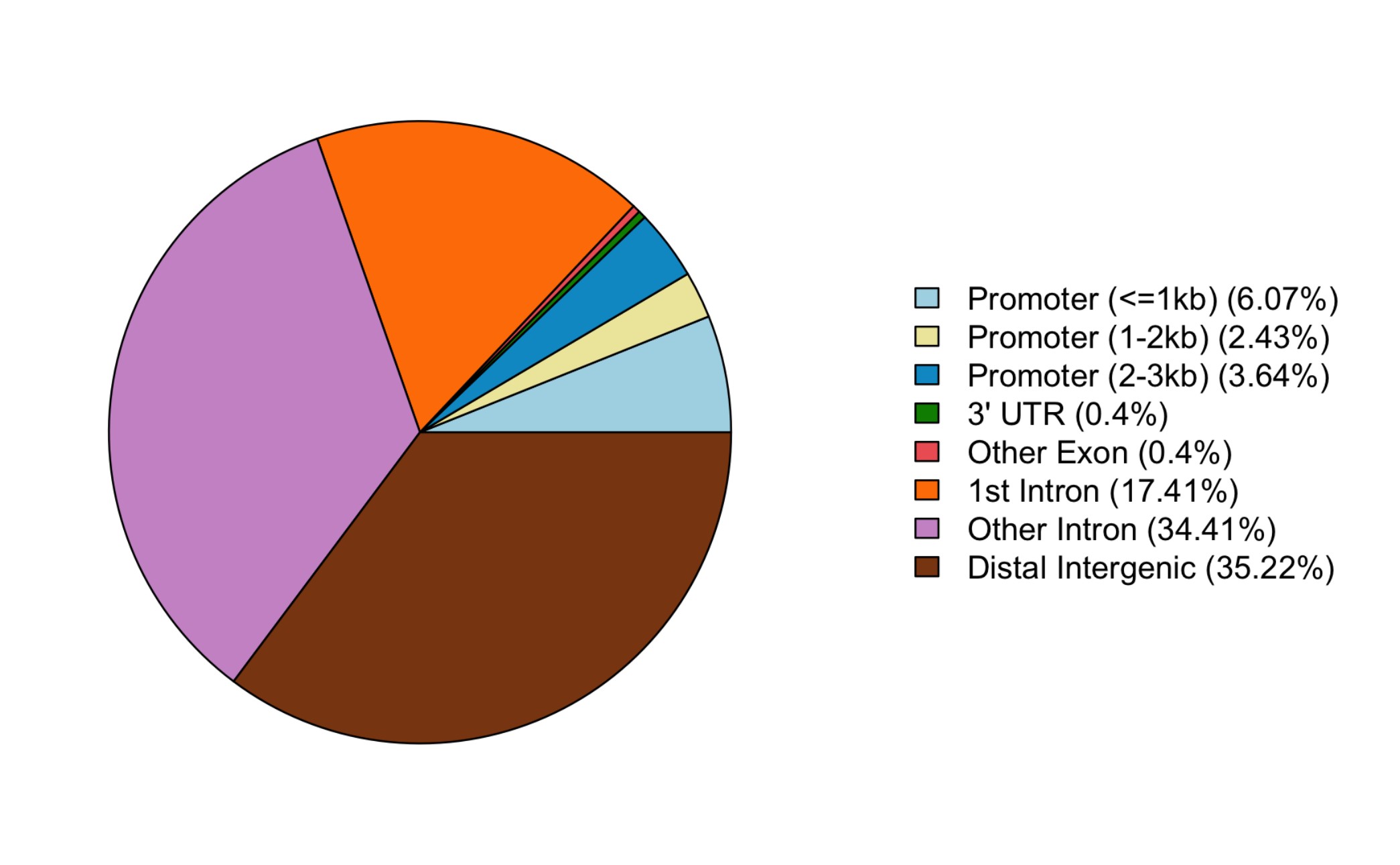

Genomic annotation of RTE loci that drive the association with sex. The pie plot was generated using ChIPseeker v1.26.2 R package.

**Fig. S6.** **Pathway enrichment analysis using proxy genes of sex-related RTE loci.**
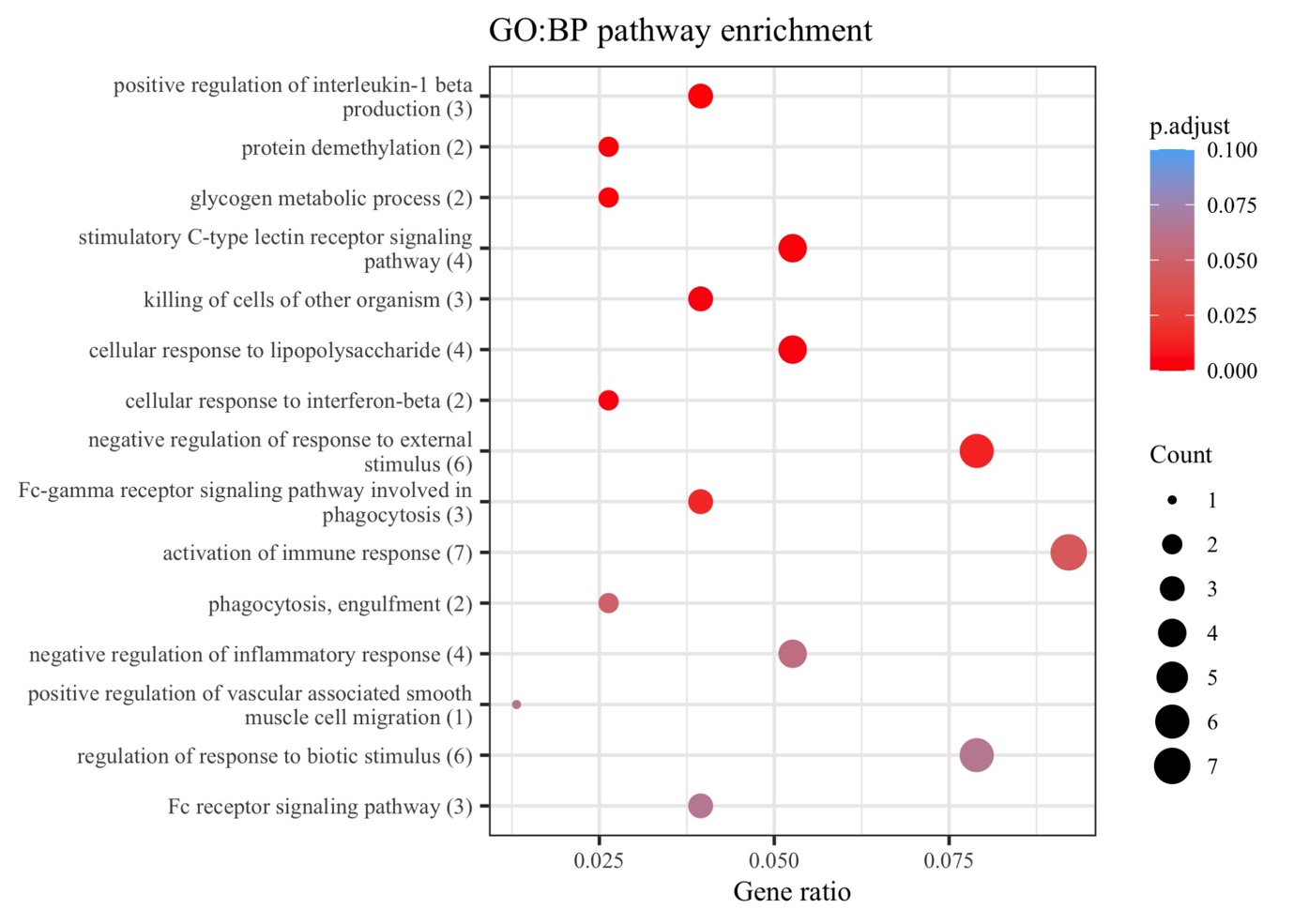

Overrepresentation analysis through Gene Ontology: Biological Process. The analysis was conducted on proxy genes of sex-related RTE loci. Only the top 15 terms are displayed. P-values were adjusted for false discovery rate.

**Fig. S7. Expression levels of MER54B subfamily in participants with low and high RTE repressor score.**

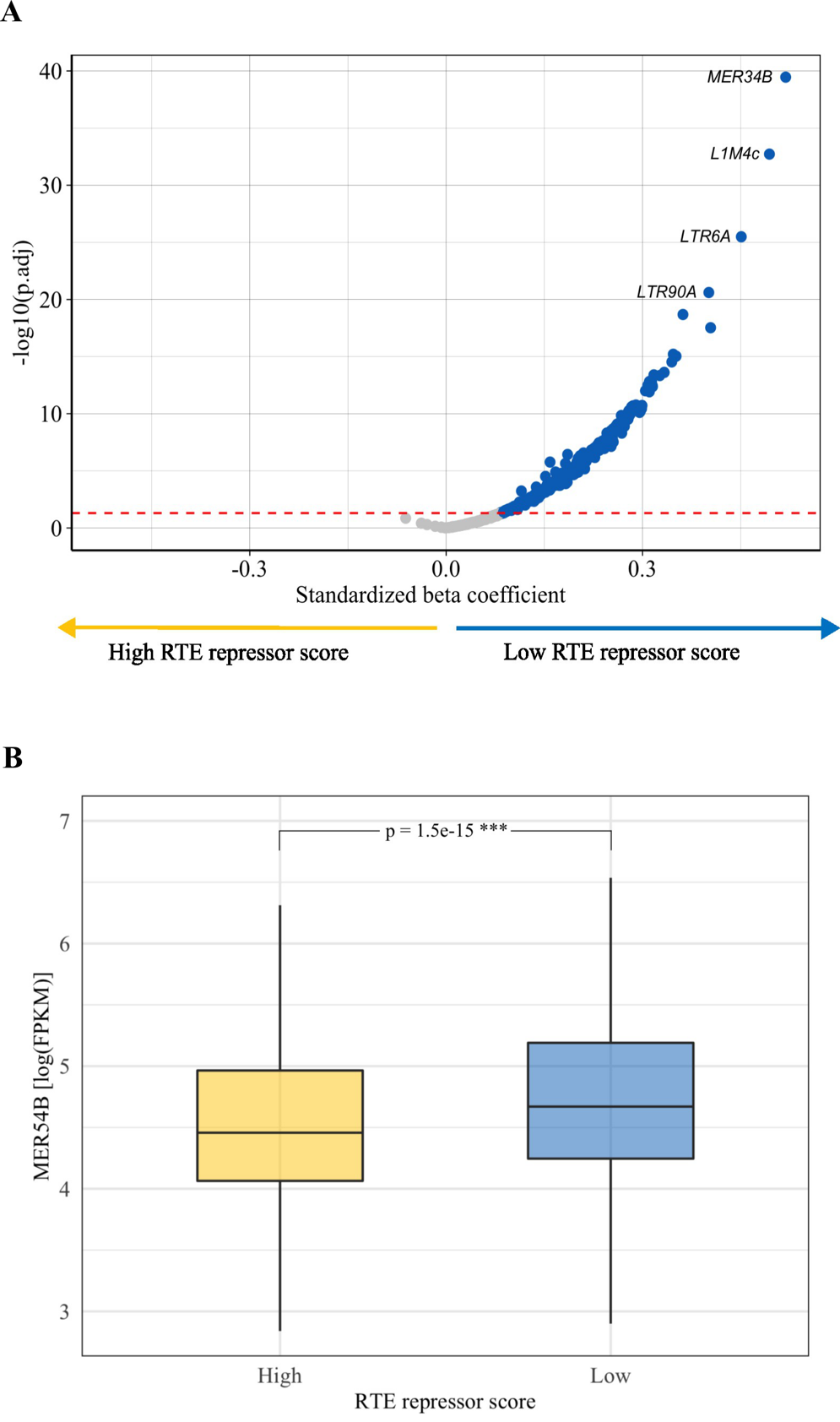

**(A)** Volcano plot showing the differences in mean expression of RTE subfamilies between the low and high RTE repressor score groups. The two groups have been defined based on the median of RTE repressor score. Each dot represents a RTE subfamily. The red line indicates the threshold for statistical significance, set at FDR < 0.05. *Model*: RTE subfamily_i_ _~_ intercept + RTE repressor score group + age + sex + smoking + batch + blood cell composition. **(B)** Box plot showing the expression levels of MER54B subfamily in participants with low *vs.* high RTE repressor score, compared statistically through an unpaired t-test.

Abbreviations: FPKM = fragments per kilobase transcript per million reads. ***p-value < 0.01
